# Chromosome X-Wide Common Variant Association Study (XWAS) in Autism Spectrum Disorder

**DOI:** 10.1101/2024.07.18.24310640

**Authors:** Marla Mendes, Desmond Zeya Chen, Worrawat Engchuan, Thiago Peixoto Leal, Bhooma Thiruvahindrapuram, Brett Trost, Jennifer L. Howe, Giovanna Pellecchia, Thomas Nalpathamkalam, Roumiana Alexandrova, Nelson Bautista Salazar, Ethan Alexander McKee, Natalia Rivera Alfaro, Meng-Chuan Lai, Sara Bandres-Ciga, Delnaz Roshandel, Clarrisa A. Bradley, Evdokia Anagnostou, Lei Sun, Stephen W. Scherer

## Abstract

Autism Spectrum Disorder (ASD) displays a notable male bias in prevalence. Research into rare (<0.1) genetic variants on the X chromosome has implicated over 20 genes in ASD pathogenesis, such as *MECP2*, *DDX3X*, and *DMD*. The "female protective effect" in ASD suggests that females may require a higher genetic burden to manifest similar symptoms as males, yet the mechanisms remain unclear. Despite technological advances in genomics, the complexity of the biological nature of sex chromosomes leave them underrepresented in genome-wide studies. Here, we conducted an X chromosome-wide association study (XWAS) using whole-genome sequencing data from 6,873 individuals with ASD (82% males) across Autism Speaks MSSNG, Simons Simplex Cohort SSC, and Simons Foundation Powering Autism Research SPARK, alongside 8,981 population controls (43% males). We analyzed 418,652 X-chromosome variants, identifying 59 associated with ASD (p-values 7.9×10⁻⁶ to 1.51×10⁻⁵), surpassing Bonferroni-corrected thresholds. Key findings include significant regions on chrXp22.2 (lead SNP=rs12687599, p=3.57×10⁻⁷) harboring *ASB9*/*ASB11*, and another encompassing *DDX53/PTCHD1-AS* long non-coding RNA (lead SNP=rs5926125, p=9.47×10⁻⁶). When mapping genes within 10kb of the 59 most significantly associated SNPs, 91 genes were found, 17 of which yielded association with ASD (*GRPR*, *AP1S2*, *DDX53*, *HDAC8*, *PCDH19*, *PTCHD1*, *PCDH11X*, *PTCHD1-AS*, *DMD*, *SYAP1*, *CNKSR2*, *GLRA2*, *OFD1*, *CDKL5*, *GPRASP2*, *NXF5*, *SH3KBP1*). *FGF13* emerged as a novel X-linked ASD candidate gene, highlighted by sex-specific differences in minor allele frequencies. These results reveal significant new insights into X chromosome biology in ASD, confirming and nominating genes and pathways for further investigation.

## 1. Introduction

Autism Spectrum Disorder (ASD [MIM 209850]) is a neurodevelopmental condition defined by social communication atypicalities, restrictive interests and repetitive sensory–motor behaviors. It is diagnosed in ∼1% of the population worldwide^1,2^, with a 3-4:1 male:female prevalence ratio^3,4^.

This difference may have demographic and social components, with one example being that some autistic traits, such as restricted interests, may be more normalized in females compared with male individuals, and consequently ASD could be underdiagnosed^5^. However, there is evidence for a significant biological influence on the sex-differential likelihood of ASD^6–11^. For example, females with neurodevelopmental disorders, including ASD, tend to have an excess of deleterious autosomal copy number variants (CNVs), and deleterious autosomal single-nucleotide variants (SNVs)^6,7,10–13^. Variation in steroid hormones and differential gene expression in males and females may also influence ASD likelihood and characteristics^8^. Another consideration, which may be influenced by the afore-mentioned observations, is that in a family with a son having ASD, the likelihood of a female sibling also being affected is 4.2%, a number that increases to 12.9% if the sib is male^14^.

Collectively the evidence may suggest a hypothetical "female protective effect" whereby females require a quantitatively greater etiologic load than males to exhibit the same degree of clinical presentation of ASD^15–17^. The sex ratio contribution approaches 1:1 when considering *de novo* mutations affecting presumed ‘penetrant’ autosomal genes and copy number variants (CNVs) in ASD and other neurodevelopmental conditions^18–20^. However, some studies suggest that the etiology of ASD includes qualitative sex differences, particularly involving genetic variations on the X chromosome.^21^. Sex hormones, known influencers of typical male and female brain development^22^, may also contribute to sex-varied penetrance in ASD^23^. For example, a surge of testosterone in the male fetus, combined with XY chromosomal determinants, may impact the neuroimmune system, affecting dendritic arborization^24^ and the number of microglia and neurons^25^, hence contribute to the sex-difference biology of ASD.

Currently, there are 23 SFARI^26^ score 1 and 36 SFARI^26^ score 2 genes with evidence to be involved in ASD mapping to the X chromosome^26^. Nine of these reach a sufficient “Evaluation of Autism Gene Link Evidence (EAGLE)” score to be considered definitively involved in more narrowingly-defined ASD^27^ (the SFARI and EAGLE genes are often used in diagnostic testing panels for ASD)^28^. Since upwards of 75% of genome-wide studies do not consider rare or common variants (including polygenic score analysis) on the sex chromosomes in their analysis ^29^, it is anticipated there are additional gene loci to be validated and others to be discovered (Table S1 summarizes the published genome-wide manuscripts examining the X-chromosome). One study has attempted a genetic association test for ASD using common variants on the X chromosome^30^, finding *TBL1X* as a candidate locus (Table S1).

There are, however, complications in studying the X chromosome as it has a lower genetic diversity compared to the autosomes, because, apart from the small pseudoautosomal region, this genomic region does not recombine in males^29^. Thus, the X chromosome can be more sensitive to evolutionary events, such as sex-bias admixture, bottlenecks and natural selection, and it can have different mutation rates from autosomes^31^. Moreover, in females, the X-inactivation phenomenon can occur where a random X chromosome copy may be inactive (i.e. X chromosome dosage compensation)^29,31,32^. The issue of 50% reduced X-chromosome coverage in males (46XY) in microarray and sequencing experiments has also led to the understudy of this important sex chromosome^29^.

Recent development, however, now enables more robust X-wide association studies (XWAS) by dealing with X-specific quality control, statistical tests stratified by sex, estimation of significant thresholds, and accounting for the potential heterogeneity of allelic effect between males and females and chromosome inactivation bias ^29,33^.

Here, we conducted a comprehensive XWAS of ASD from 6,873 ASD individuals (5,639 males and 1,234 females) sourced from three different whole-genome sequencing (WGS) datasets, alongside 8,981 control individuals (3,911 males and 5,070 females), from two additional datasets (Figure 1, Figure S1).

**Figure 1:**
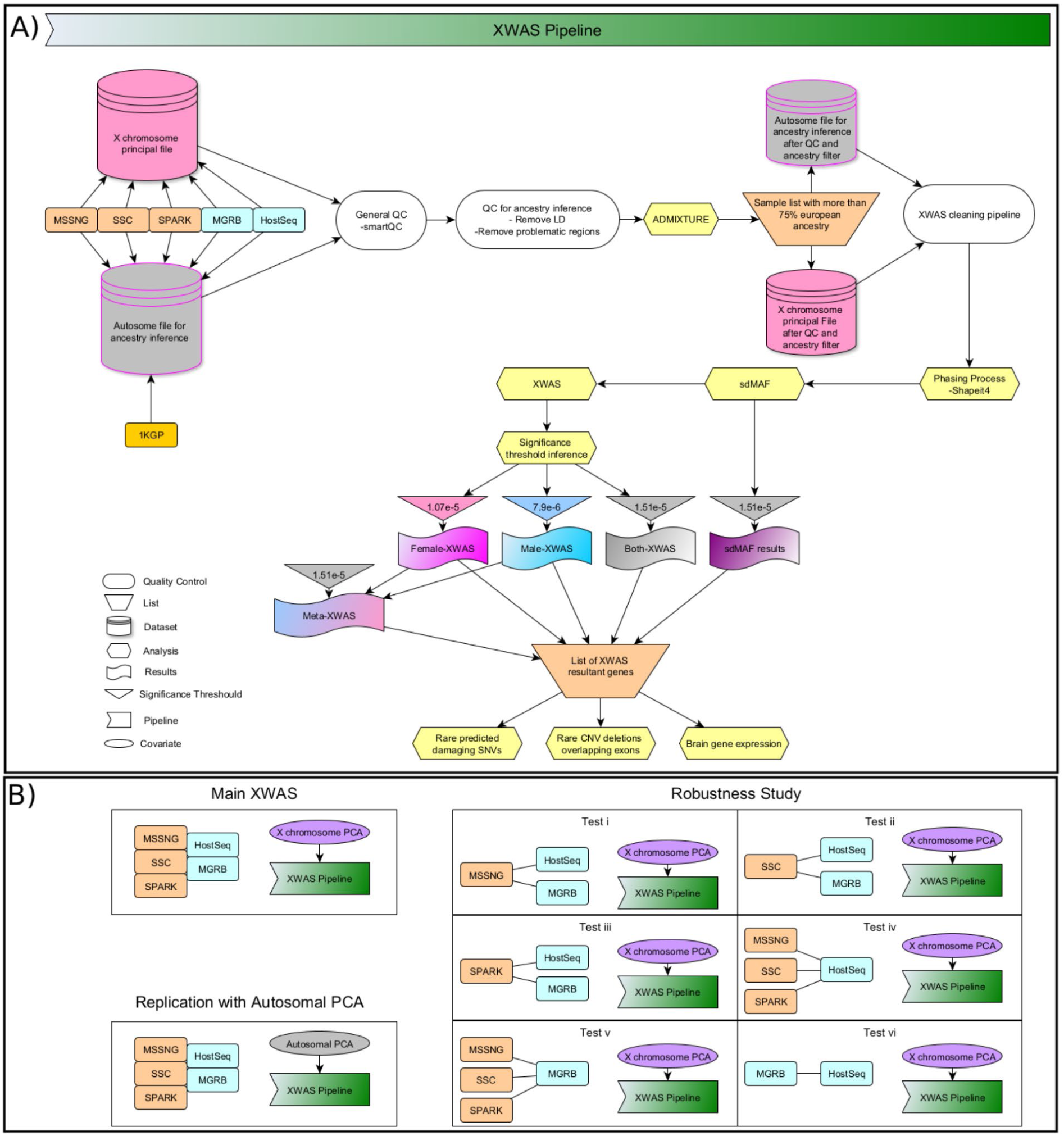
XWAS workflow. A) Outline of the XWAS pipeline detailing data sources including MSSNG (Autism Speaks), SSC (Simons Simplex Cohort), SPARK (Simons Foundation Powering Autism Research), 1KGP (1000 Genome Project), HostSeq (The Host Genome Sequencing Initiative), and MGRB (Medical Genome Reference Bank). The significance threshold was determined using Bonferroni correction, individually calculated for the Male-XWAS, Female-XWAS, and Both-XWAS approaches. For Meta-XWAS, we used the threshold inferred from the Both-XWAS result. B) Replication and robustness studies conducted.

## 2. Material and methods

### 2.1 Database

#### 2.1.1. ASD Datasets

The Autism Speaks MSSNG resource^34,35^ is a dataset of genetic and phenotype information from individuals diagnosed with ASD as well as members of their families ^34,35^. The affected individuals were diagnosed according to the Diagnostic and Statistical Manual of Mental Disorders (DSM)^36^, also supported in many individuals by the Autism Diagnostic Interview-Revised (ADI-R)^37,38^ and/or the Autism Diagnostic Observation Schedule (ADOS) ^39,40^. The Province of Ontario Neurodevelopmental Network (POND) is part of MSSNG and continues to contribute with new data. We used data from 9,621 individuals for the analysis done here.

The Simons Simplex Collection (SSC) includes WGS data from approximately 2,600 ASD simplex families (one affected child plus unaffected parents and siblings)^41^. The ASD diagnoses were performed following the University of Michigan Autism and Communication Disorders Center guidance to guarantee uniformity across the 12 university-affiliated research clinics involved in this initiative. We used 9,209 ASD participants from SSC in this analysis. Also from SFARI^42^, the SPARK data (Simons Foundation Powering Autism Research) is an autism research initiative that includes both WES (Whole Exome Sequence) and WGS data from US individuals, besides behaviour and phenotypic information. We used WGS from 12,519 individuals for the X-chromosome analysis.

#### 2.1.2. Population/Control Datasets

For ancestry inference, we used genetic information from 3,202 samples from 1000 Genomes Project of five different ancestries (Africans, Americans, East Asians, Europeans and South Asians). For this, we used the high-coverage 2020 version released by the New York Genome Center (NYGC) (https://www.internationalgenome.org/data-portal/data-collection/30x-grch38)^43^.

As ASD-controls we used data from 2,561 samples from the Medical Genome Reference Bank (MGRB)^44^, which is a WGS dataset from ∼4,000 healthy, elderly Australians^44^. The MGRB dataset includes most individuals of European ancestry but does not exclude samples from different genetic backgrounds. We also used 9,802 samples from the Host Genome Sequencing Initiative (HostSeq)^45^ which is a collection of 14 Canadian research studies examining responses to COVID-19.

### 2.2 Quality Control

#### 2.2.1 Autosomes

After selecting biallelic variants we used the SmartQC software (https://github.com/ldgh/MosaiQC-public) to perform the basic control quality steps to remove: (i) variants with the chromosome notation equal to “0”, (ii) remove variants with duplicated IDs, (iii) remove variants and samples with missing data greater than 10% (plink --geno 0.1; plink --mind 0.1), (iv) impute sex codes using SNP data through PLINK’s ’--impute-sex --check-sex’ functionality. (v) remove A|T and C|G variants (ambiguous SNPs), (vi) remove 100% heterozygous variants (plink --hardy) and (vii) annotate the variants for dbSNP ID and LiftOver for hg38 if necessary.

Using plink --bmerge, we merged the data from MSSNG, SSC, SPARK, MGRB, HostSeq. The merged file had a total of 22,242 samples and 1,407,803 variants (Figure 1, Figure S1).

For the XWAS analysis using Principal Components (PCs) based on the autosomal information as covariates for logistic regression, we cleaned our data based on the pipeline of Leal *et al* (2023)^46^ (https://github.com/MataLabCCF/XWAS) in the merged file with MSSNG, SSC, SPARK, MGRB, and HostSeq. This cleaning pipeline adds the following steps; (i) removal of monomorphic SNPs, or those located in structural variants, using the list of SNPs located in structural variants from Le Guen *et al*. (2021)^32^ created using Tri-Typer^47^. (ii) remove of potential probe sites using gnomAD^48^, and (iii) relationship control using KING^49^ to calculate the kinship coefficient and NAToRA^50^ to remove samples with relatedness closer than second degree. After this XWAS cleaning pipeline the autosomal file had 1,075,065 SNPs and 21,089 samples (Figure S1).

The final XWAS analysis was restricted to individuals with more than 75% European ancestry. To achieve this, we conducted an ancestry check utilizing ADMIXTURE software^51^ with five clusters. The reference populations included Europeans, Africans, East Asians, South Asians, and Americans from the 1000 Genomes Project (1KGP)^52^. After merging our XWAS data with samples from the 1000 Genomes Project (1KGP), which underwent the same quality control process, we obtained a dataset containing 24,291 samples (Figure S1). To enhance data quality for ancestry inference, we conducted a filtering step to exclude variants exhibiting high levels of Linkage Disequilibrium (LD), using the command ’plink --indep-pairwise 100 10 0.1’. Additionally, variants located in regions known to be under recent selection were removed from the dataset^53–55^. We then ran ADMIXTURE using a total of 131,291 SNPs.

#### 2.2.2 X Chromosome

After completing the general quality control steps described in section 2.2.1, we separated the variants on the X chromosome (coded as chromosome 23 in PLINK) from those in the pseudoautosomal regions (coded as chromosome 25 in PLINK). This separation was based on a dbSNP reference file. We also applied the XWAS cleaning pipeline (Figure 1)^46^ for the X chromosome, which includes; (i) selecting the remaining individuals from the autosomal cleaning process, including samples without relatedness greater than second degree and samples with more than 75% of European ancestry, (ii) removal of SNPs following the same parameters used for the autosomes, besides SNPs with differential missingness between ASD individuals and controls with p-values lower than 10^-5^, (iii) removal of SNPs with differential missingness between males and females with p-values lower than 10^-5^, (v) heterozygous SNPs found in males were assigned as missing data. For the XWAS logistic regression we used a final of 418,652 X chromosomal variants and 15,499 samples (Figure S1).

### 2.3. XWAS

After data cleaning, we conducted the XWAS analysis using two input files. The first file contained autosomal data with 1,075,065 variants, intended for principal component inferences to be used as covariates in the XWAS logistic regression. The second file consisted of X chromosome data with 418,652 variants. Both files contained data from the same 15,854 samples. Among these samples, 9,550 were male (3,911 controls and 5,639 ASD individuals), and 6,304 were females (5,070 controls and 1,234 ASD individuals). The principal component inference was done with the GENESIS package stratified by sex (one PCA for males and one for females), where all samples with standard deviation greater than three from major clusters were defined as being outliers and removed from subsequent analyses. For the primary XWAS analysis, we utilized X chromosome data for principal component analysis (PCA). Additionally, we conducted a replication analysis using data from autosomal chromosomes. The resulting 10 PCs from males only and females only were employed as covariates for Male-XWAS and Female-XWAS, respectively. Non-outlier males and females were merged to create both datasets. Subsequently, this new dataset underwent another PCA, where outliers were detected and excluded based on the same parameters. The 10 resulting PCs from this process were used as covariates in “Both-XWAS” (Figure 1).

The final regression analysis was performed using logistic regression in PLINK2 ^56^ (--glm) for three approaches (Figure 1); (i) Male-XWAS: Based on 5,639 ASD male individuals and 3,911 male controls. This approach used the 10 top PCs from males as covariates; (ii) Female-XWAS: Based on 1,234 ASD female individuals and 5,070 female controls. This approach used the 10 top PCs from females as covariates; and (iii) Both-XWAS: Based on 6,873 ASD individuals and 8,981 controls. This approach used the 10 top PCs from both and sex as covariates.

We also performed a meta-analysis from the sex-stratified results (Male-XWAS and Female-XWAS) implemented on GWAMA^57,58^. This result incorporates the "gender_heterogeneity_p-value," which is derived from assessing heterogeneity between sex-specific allelic effects. This result incorporates the "gender_heterogeneity_p-value," which is derived from assessing heterogeneity between sex-specific allelic effects using one degree of freedom. This test involved analyzing males and females separately in each XWAS. It entailed obtaining male- and female-specific allelic effect estimates in a fixed-effects meta-analysis, followed by testing for heterogeneity between the sexes ^58^.

### 2.4 X-Chromosome Significance Threshold

Given that our association tests are conducted on a single chromosome, the number of effective tests performed is lower compared to a genome-wide analysis. Typically, in genome-wide analyses, the significance threshold is set at p-value < 5×10^-8^. To determine an appropriate significance threshold for our XWAS analysis, we applied a Bonferroni correction by dividing 0.05 by the number of effective tests ^59^.

The number of effective tests (N_eff_) was calculated by dividing the squared number of variants by the sum of the R^2^ correlation coefficients between all variants present in the dataset ^46^:

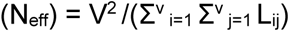

V= Total number of variants

L = the R² correlation coefficient between all variants (V) present in the datasets (L is a matrix with size V by V); i and j = Matrix indexes.

To generate the R2 matrix among all variants in our dataset, we utilized the command ’plink -

-r2 square gz yes-really’. The sum of our corresponding matrix was: Female: 37441288.90; Male: 27721944.23 and Both: 53006792.62. Thus, the respective number of effective tests (N_eff_): Female: 418,652^2^/37441288.90 = 4,681.18; Male: 418,652^2^/ 27721944.23 = 6,322.40; Both: 418,652^2^/ 53006792.62 = 3,306.54 with the final significance threshold being; Female: 0.05/4,681.18 = 1.07×10^-5^; Male: 0.05/6,322.40 = 7.9×10^-6^; Both: 0.05/3,306.54 = 1.51×10^-5^.

### 2.5. sdMAF

Sex differences in allele frequencies were analyzed with the sdMAF software ^60,61^. We initially split the pseudoautosomal regions (PAR) with the PLINK^62^ --split-par hg38 command. Since PLINK was not able to properly handle male homozygous in the bed file and simply assigned them all to missing, we bypassed the problem by changing the chromosome code to 22 prior to generating genotype counts. The chromosome number in the ‘gcount’ file was then re-coded back to 23 and subsequently pipelined into the sdMAF software as suggested by the sdMAF documentation^61^. To select the significant sdMAF results, we utilized the same conservative, Bonferroni-corrected significance level for XWAS-Both analysis (1.51×10^-5^), given that we are testing the same number of SNPs.

### 2.6 Rare variant analysis

We further investigated the impact of rare genetic variations inside the candidate regions identified from the XWAS analysis by comparing the frequency of rare predicted damaging single nucleotide variants (SNVs; gnomAD frequency <0.1%), insertion and deletions smaller than 50bp (indels; gnomAD frequency <0.1%), and exonic copy number deletions (CNV deletions; gnomAD frequency <1%) impacting genes between ASD-probands and family members.

The initial reads were aligned to the GRCh38 human genome reference. Small variants (SNVs and Indels) and CNVs were called using GATK and *in-house* CNV calling pipeline, respectively^63^. Standard output files were generated, including CRAMs for alignment, and VCFs for small variants, and CNVs. Per sample analysis metrics were also generated. The small variant calls were annotated using an ANNOVAR-based pipeline^64^. Using an in-house script, we filtered high quality small variants that were found in less than 0.1% of gnomAD samples. We then selected only damaging small variants if they result in a stop gain or a frameshift, or, they are nonsynonymous SNVs predicted to be damaging by four different in-silico tools (i.e., sift_score^65^ <=0.05, polyphen_score^66^>=0.9, mt_score^67^>=0.5, and CADD_phred^68^ >= 15). For this SNVs analysis, besides the WGS data previous described (session 2.1.1), we also used whole exome data (WES) from SPARK, given a final number of 47,840 ASD-probands (79% males), 19,820 ASD-unaffected siblings (47% males), and 63,692 ASD-parents (40% fathers).

The deletions were detected using a previously described read depth-based pipeline^34,63^. We only considered high-quality deletions, which were tagged based on the following criteria; (i) length >= 5kb, ii. called by both ERDS^69^ and CNVnator^70^ with at least 50% reciprocally overlapped in length, (ii) having < 70% of its length overlap with repetitive or low complexity regions of the genome (i.e., telomere, centromere, and segmental duplications), and (iii). for the X chromosomal calls in males, CNVs in PAR were filtered out. For the CNV comparison we only used WGS data, and we also included data from the new MSSNG release (MSSNGdb7), resulting in a total of 9,691 ASD-probands (82% males), 5,591 ASD-unaffected siblings (38% males) and 17,470 ASD-parents (50% fathers).

For both small variants and deletions, independently, we performed an association analysis using a conditional logistic regression stratifying the test by the family. For sex-combined analysis, we also used sex as covariates.

### 2.7 Brain gene Expression Analysis

Exon-averaged gene expression data were obtained from BrainSpan (Allen Brain Atlas)^71^. With this microarray data, we further applied quantile normalization and standardization across both genes and samples for the comparative analysis. Subsequently, we generated a brain map plot wherein colors ranging from blue (indicating downregulation) to red (indicating upregulation) denote the average expression levels of the selected genes within each brain region. This visualization was created for five developmental stages: Early Fetal (less than 16 weeks), Late Fetal (more than 16 weeks to birth), Early Childhood (birth to three years old), Childhood/Teenage (three years to 20 years), and Adulthood (more than 20 years).

## 3. Results

### 3.1 Association Test

After performing the four different XWAS tests (Figure 1), which included sex-stratified tests (Male-XWAS and Female-XWAS), sex-combined mega-analysis (Both-XWAS), and meta-analysis (Meta-XWAS), we identified 59 variants as significant in at least one of the four approaches (Table S2). These variants correspond to a total of 20 risk loci, encompassing 23 genes with variants in high linkage disequilibrium (r^2 > 0.7) with the lead SNP (Table 1). The genomic loci detected from the four XWAS approaches utilized are shown in Table 1. Among these, 42 were found uniquely by a unitary XWAS approach: 27 in the Male-XWAS, five in the Female-XWAS, one in the Both-XWAS (performed with males and females together, using sex as a covariate), and nine in the Meta-XWAS (a meta-analysis of Male-XWAS and Female-XWAS results using GWAMA^57^ software, because it includes a "meta-analysis using sex-differentiated and sex heterogeneity"^57^). Additionally, 17 variants showed significant p-values in more than one test (Table S2). Each test underwent visual inspection via histograms and QQ plots, revealing no distortions as indicated by the genomic inflation factor (λ=0.928 - 1.036), which measures systematic bias in the statistical test (Figure S2, Figure 2). Among the 59 variants, 30 exhibit a “gender heterogeneity p-value” (test for heterogeneity between sexes with one degree of freedom)^58^ below 0.05, all of them in the sex stratified approaches (26 in the Male-XWAS and four in the Female-XWAS), suggesting significant differences in allelic effects between males and females for these variants. Notably, two of these variants, identified in the Male-XWAS within the *ASB11* gene, attained a “gender heterogeneity p-value” of less than 9×10^−5^. In the presence of heterogeneity in allelic effects between the sexes, a loss of power for sex-combined association tests can occur if the allele has opposite direction of effect in the other sex^58^. This type of biological phenomena may explain why variants are not detected in Both-XWAS and Meta-XWAS.

**Figure 2.**
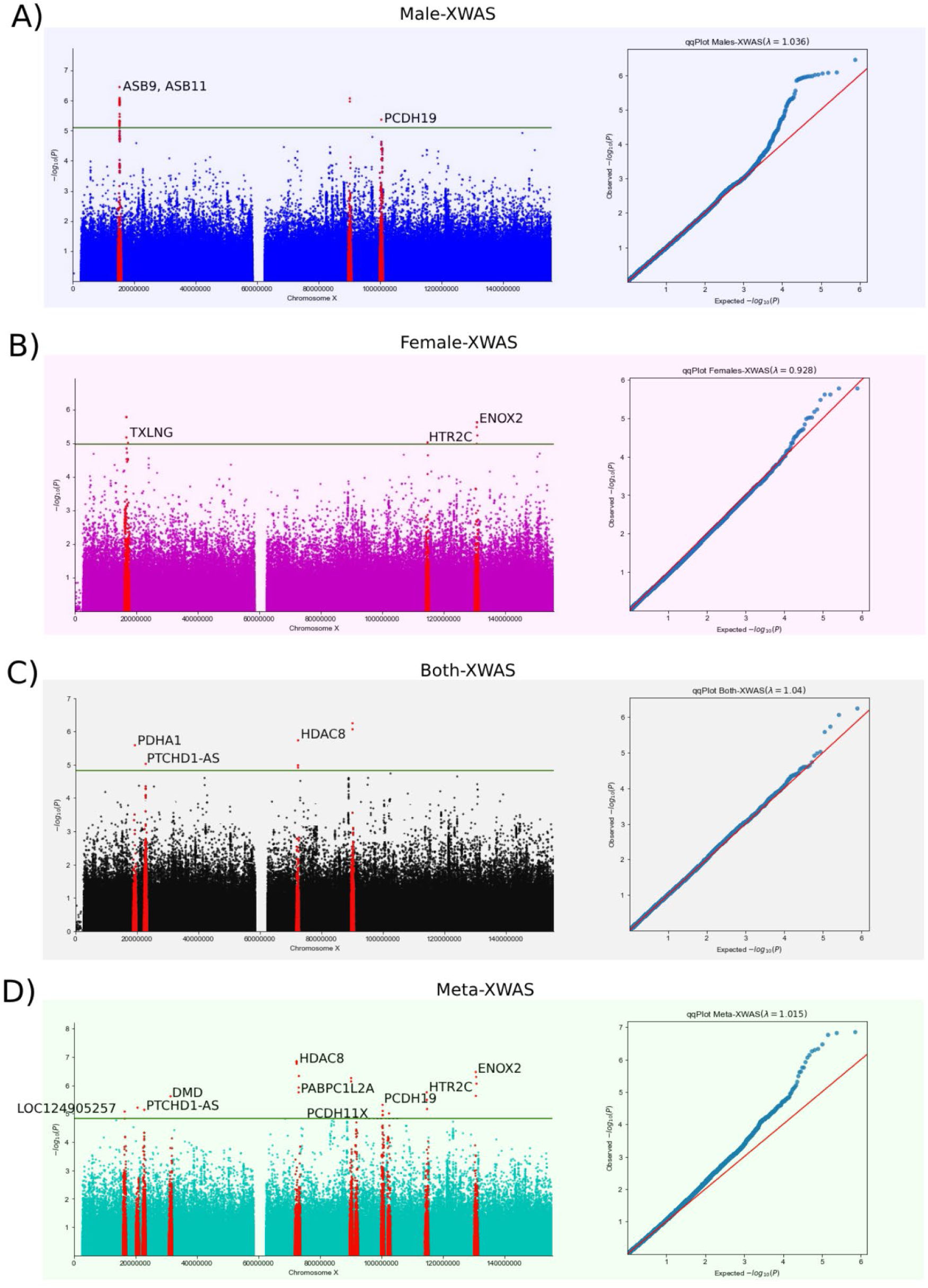
ASD-XWAS manhattan and qq plots. Each panel shows a Manhattan plot on the left part and qqPlot on the right part. The graphs result from XWAS testing using 6,873 ASD individuals (5,639 males and 1,234 females) and 8,981 controls (3,911 males and 5,070 females) with a total of 418,652 X chromosomal variants originated from WGS data (46 variants in PAR regions) for (A) Male-XWAS, B) Female-XWAS, C) Both-XWAS, and D) the Meta-XWAS, a meta-analysis from the sex stratified approaches implemented on GWAMA^57^.

**Table 1.** Genomic Risk Loci. Genomic loci detected from four XWAS analyses (Male-XWAS, Female-XWAS, Both-XWAS, Meta-XWAS). The unique ID as well as the p-value refer to the lead SNPs specified. The 23 genes in the last column are within the gene locus, and encompass variants exhibiting strong linkage disequilibrium with the lead SNP (r2 > 0.7).

| XWAS approach | Genomic Locus | UniqueID | Lead SNPs | p | start_hg37 | end_hg37 | start_hg38 | end_hg38 | Size_hg38 | Genes |
| --- | --- | --- | --- | --- | --- | --- | --- | --- | --- | --- |
| Males | 1_Male-XWAS | 23:15283903:A:C | rs12687599 | 3.57E-07 | 15274671 | 15366179 | 15256549 | 15348057 | 91508 | ASB9, ASB11, PIGA |
| Males | 2_Male-XWAS | 23:89417731:C:T | rs148591304 | 1.09E-06 | 89417731 | 89426923 | 90162732 | 90171924 | 9192 | Intergenic |
| Males | 3_Male-XWAS | 23:99660751:C:T | rs12835197 | 4.33E-06 | 99631253 | 99686238 | 100376255 | 100431240 | 54985 | PCDH19 |
| Female | 1_Female-XWAS | 23:16787511:C:T | rs185190849 | 1.68E-06 | 16683001 | 17354197 | 16664878 | 17336074 | 671196 | TXLNG, CTPS2, REPS2 |
| Female | 2_Female-XWAS | 23:113992014:A:G | rs191071511 | 9.58E-06 | 113958523 | 113992014 | 114724114 | 114757574 | 33460 | HTR2C |
| Female | 3_Female-XWAS | 23:129957059:C:T | rs186814690 | 2.41E-06 | 129769645 | 130270300 | 130635671 | 131136326 | 500655 | ENOX2 |
| Both | 1_Both-XWAS | 23:19374946:C:T | rs767542284 | 2.61E-06 | 19215082 | 20143856 | 19196964 | 20125738 | 928774 | PDHA1 |
| Both | 2_Both-XWAS | 23:22885946:A:G | rs5926125 | 9.47E-06 | 22854092 | 22893611 | 22835975 | 22875494 | 39519 | PTCHD1-AS |
| Both | 3_Both-XWAS | 23:71657264:A:G | rs5958792 | 1.85E-06 | 71657264 | 71657264 | 72437414 | 72437414 | 0 | HDAC8 |
| Both | 4_Both-XWAS | 23:89417731:C:T | rs148591304 | 5.69E-07 | 89417731 | 89426923 | 90162732 | 90171924 | 9192 | Intergenic |
| Meta | 1_Meta-XWAS | 23:16522448:C:T | rs111827716 | 8.40E-06 | 16487766 | 16523991 | 16469643 | 16505868 | 36225 | Intergenic |
| Meta | 2_Meta-XWAS | 23:20723519:A:G | rs776360992 | 6.15E-06 | 20723519 | 20748993 | 20705401 | 20730875 | 25474 | LOC124905257 |
| Meta | 3_Meta-XWAS | 23:22885946:A:G | rs5926125 | 7.30E-06 | 22854092 | 22893611 | 22835975 | 22875494 | 39519 | PTCHD1-AS |
| Meta | 4_Meta-XWAS | 23:31460970:A:G | rs139802025 | 2.44E-06 | 31460970 | 31460970 | 31442853 | 31442853 | 0 | DMD |
| Meta | 5_Meta-XWAS | 23:71623954:G:T | rs73218354 | 1.42E-07 | 71623954 | 72600433 | 72404104 | 73380597 | 976493 | HDAC8, PABPC1L2A |
| Meta | 6_Meta-XWAS | 23:89417731:C:T | rs148591304 | 5.53E-07 | 89417731 | 91160501 | 90162732 | 91905502 | 1742770 | PCDH11X |
| Meta | 7_Meta-XWAS | 23:99660751:C:T | rs12835197 | 4.87E-06 | 99631253 | 99686238 | 100376255 | 100431240 | 54985 | PCDH19 |
| Meta | 8_Meta-XWAS | 23:101756502:C:T | rs5945876 | 9.79E-06 | 101060265 | 101847597 | 101805292 | 102592669 | 787377 | TCP11X3P, BEX5 |
| Meta | 9_Meta-XWAS | 23:113958523:G:T | rs140894960 | 1.76E-06 | 113939197 | 114789771 | 114704781 | 115555444 | 850663 | HTR2C |
| Meta | 10_Meta-XWAS | 23:129834346:A:G | rs189525731 | 3.40E-07 | 129223355 | 130791852 | 130089380 | 131657839 | 1568459 | ENOX2 |

#### 3.1.2 Robustness Study

We performed XWAS analyses using various configurations, including one ASD dataset against all controls, as well as all ASD against each control dataset, to mitigate potential bias stemming from dataset heterogeneity and to conduct robust sanity replications, (Table S3). Consequently, we obtained XWAS results for; (i) MSSNG as cases versus HostSeq and MGRB as controls, (ii) SSC as cases versus HostSeq and MGRB as controls, (iii) SPARK as cases versus HostSeq and MGRB as controls, (iv) MSSNG, SSC, and SPARK as cases versus HostSeq as controls, (v) MSSNG, SSC, and SPARK as cases versus MGRB as controls and (vi) control versus control (sanity test; MGRB was labeled as cases and HostSeq as control).

Among the 27 variants exclusively found in males, all replication tests yielded a p-value below 0.05, except for eight variants solely in robustness test "v" (involving all case datasets versus MGRB controls). Notably, these eight variants reside within the first significant region identified in the Male-XWAS, spanning between 15.27 and 15.36 Mb. Even after excluding these eight variants, we retained 19 significant SNPs in this region with a p-value lower than 0.05 across all replication tests.

Regarding the five variants in the detected exclusive in Female-XWAS, two did not reach a p-value lower than 0.05 across all robustness tests. One of them, rs749183760 in *ENOX2*, was not captured by test "ii". Additionally, the intergenic SNP rs182249604, located within the first significant genomic region (between 16.7Mb and 17.33Mb), did not yield a p-value lower than 0.05 on test “iii”. However, even after excluding these variants, we still observe significant SNPs in this region, including variants within the *TXLN* gene.

Considering the results from the Both-XWAS replications, there is only one significant variant (rs767542284 in *PDHA1*), that also demonstrated a significant p-value in all subset (i-v) analyses; five of nine variants detected in the Meta-XWAS achieved significant p-values in all cohort tests, and these are located within *DMD*, *PABPC1L2A* and *PCDH11X*. When comparing the significant variants detected in both Meta-XWAS and Both-XWAS (8 variants located on *PTCHD1-AS*, *HDAC8*, and *LOC124905257* genes), we replicated five results across all cohort tests (i to v). Notably, the variants that did not reach a significant p-value in all tests include two variants in the *HDAC8* gene (rs5958792, rs73218354) and one intergenic variant (rs5981334), all of which were not replicated only in test "iii" (SPARK versus all control cohorts).

All six results identified in both the Female-XWAS and another XWAS (Meta-XWAS, Both-XWAS) were situated within two different genes, *ENOX2* and *HTR2C*. None of these variants achieved a p-value < 0.05 in test ii (SSC versus all controls). Three significant variants were detected in both the Male-XWAS and Meta-XWAS. All three variants had significant p-values in all tests except for one variant (rs12835197 - *PCDH19*) in test "v" (All cases versus MGRB).

We performed a sanity check employing logistic regression, where controls were compared against controls (Test “vi”), using MGRB as cases and Hostseq as controls. To fortify the reliability of our findings, we assessed whether our candidate variants yielded non-significant p-values (≥0.05) in this sanity test as well. At least one variant in the genes *ASB9* and *ASB11* from Male-XWAS analysis meet the criteria of the sanity test. Hence, we retained both genes in the final results. In the Female-XWAS results, three out of the five detected variants failed to pass the sanity test, resulting in only the ENOX2 gene being included among the final genes.

#### 3.1.3 XWAS replication using Autosomal Principal Components as covariates

Our principal component analysis (PCA) focused solely on the X chromosome due to its unique biological features (see Methods section 2.3). Therefore, the top 10 PCs were then considered as covariates^46^. We also implemented XWAS with autosomal PCs to assess the generalizability of findings (Table S4, Figure S3 and Figure S4). The modified model revealed a total of 58 significant loci spanning over 12 genes: *ASB9*, *ASB11*, *PIGA*, *PCDH19*, *TXLNG*, *HTR2C*, *ENOX2*, *PDHA1*, *PTCHD1-AS*, *DMD*, *HDAC8*, and *PABPC1L2A* (Table 2); 11 of these overlap with the genes detected by the primary analysis using X chromosome-only PCs (Figure S4). The *PIGA* gene was identified exclusively with the autosomal PC model, noting it is located in proximity to *ASB11*(3.8kb) and *ASB9* (48.9kb), which were detected in the Male-XWAS results using the X chromosome PCs. Among the 14 genes discovered by the XWAS using the X chromosome-only PCs only X, *LOC124905257* and *PCDH11X* were not present when using autosomal PCs in the XWAS.

**Table 2.** Significantly associated ASD genes based on our main XWAS and sdMAF results. The values in red are the p-values considered significant based on the specific Bonferroni corrections (Males: 7.9×10^-6^, Females: 1.07×10^-5^, Both: 1.51×10^-5^).

| Gene | Dataset | LeadSNP | P_Meta | P_Males | P_Females | P_Both | Other Evidences |
| --- | --- | --- | --- | --- | --- | --- | --- |
| ASB9 | Males | rs12687599 | 5.74E-04 | 3.57E-07 | 6.79E-01 | 5.56E-04 | Robustness test, Replication using Autosomal PCs, rare variants detected |
| ASB11 | Males | rs6628945 | 1.22E-03 | 8.27E-07 | 6.01E-01 | 1.12E-03 | Robustness test, Replication using Autosomal PCs, rare variants detected |
| TXLNG | Females | rs753342681 | 1.31E-03 | 5.08E-01 | 1.68E-06 | 9.01E-03 | Robustness test, Replication using Autosomal PCs, rare variants detected |
| PDHA1 | Both | rs767542284 | 3.01E-05 | 7.93E-04 | 2.62E-03 | 2.61E-06 | Robustness test, Replication using Autosomal PCs, rare variants detected |
| LOC124905257 | Meta | rs776360992 | 6.15E-06 | 2.60E-05 | 5.11E-02 | 4.29E-05 | Robustness test |
| PTCHD1-AS | Meta, Both | rs5926125 | 7.30E-06 | 1.89E-04 | 1.04E-02 | 9.47E-06 | Trost; et al (2023), Autdb, SFARI 2, Robustness test, Replication using Autosomal PCs, rare variants detected |
| DMD | Meta | rs139802025 | 2.44E-06 | 8.35E-05 | 7.82E-03 | 5.33E-04 | Trost; et al (2023), Autdb 3, SFARI S, Replication using Autosomal PCs, rare variants detected |
| HDAC8 | Meta, Both | rs73218354 | 1.42E-07 | 8.14E-05 | 3.92E-04 | 1.04E-05 | Trost; et al (2023), Autdb2, SFARI S, Robustness test, Replication using Autosomal PCs, rare variants detected |
| PABPC1L2A | Meta | rs115820229 | 1.76E-06 | 1.70E-04 | 3.09E-03 | 1.18E-04 | Robustness test, Replication using Autosomal PCs |
| PCDH11X | Meta | rs184604300 | 1.38E-05 | 2.25E-04 | 2.12E-02 | 4.92E-03 | Autdb2, SFARI 2, Robustness test, rare variants detected |
| PCDH19 | Males, Meta | rs12835197 | 4.87E-06 | 4.33E-06 | 8.30E-02 | 3.94E-04 | Autdb4, SFARI 1S, rare variants detected |
| HTR2C | Females, Meta | rs140894960 | 1.76E-06 | 7.01E-02 | 9.58E-06 | 3.95E-03 | Replication using Autosomal PCs, rare variants detected |
| ENOX2 | Females, Meta | rs186814690 | 5.06E-07 | 2.68E-02 | 2.41E-06 | 5.59E-05 | CNVfreq, Replication using Autosomal PCs, rare variants detected |
| FGF13 | sdMAF | rs555083289 | sdMAF pvalue = 1.19E-05 |  |  |  | Autdb 2, SFARI 3S, rare variants detected |

The genomic control lambdas observed in the QQ plots ranged from 0.913 to 1.119 (Figure S4). Additionally, the correlation between the XWAS results obtained from X chromosome PCs and autosomal PCs was 0.76 for males and 0.79 for females (Figure S3). Overall, the results from the main analysis are generalizable and robust.

#### 3.1.4 Annotation

All XWAS results (Male-XWAS, Female-XWAS, Both-XWAS, Meta-XWAS) were annotated using both modules of FUMA^72^: SNP2GENE and GENE2FUNC. SNP2GENE mapped the genes corresponding to the significantly associated SNPs, while GENE2FUNC annotated gene expression and gene sets from the previously mapped genes. The 59 significant associated variants were mapped (within a 10kb distance) to a total of 93 genes (Table S5). Through the gene-based test conducted using MAGMA^73^, significant associations were identified for *ASB11* (p-value = 2.87×10^-6^) in the Male-XWAS (Figure 2, Figure 3-A), where initial SNPs were mapped to 704 genes given a significance threshold defined as 7.1×10^-5^ (0.05/704). This gene was also mapped in the Female-XWAS analysis, being situated within at least 10kb distance from a significantly associated SNP. Notably, *ASB11* is located within one of the most significant Linkage Disequilibrium (LD) regions identified in the Male-XWAS results, spanning between 15.27 and 15.36Mb, which also encompasses the genes *ASB9* and *PIGA* (Genomic Locus 1-Male-XWAS; Table 1). The corresponding LocusZoom plot, along with the Combined Annotation Dependent Depletion (CADD)^68,74^ score and RegulomeDB score^75,76^ plots for this region, are presented in Figure 3-A. The CADD score assesses the deleteriousness of genetic variants, while the RegulomeDB score evaluates their functional significance, aiding in the interpretation of their potential biological effects. When considering only significant SNPs falling internal to the gene rather than within a 10kb range, 13 candidates were identified: *ASB11*, *ASB9*, *DMD*, *ENOX2*, *HDAC8*, *HTR2C*, *LOC124905257*, *PABPC1L2A*, *PCDH11X*, *PCDH19*, *PDHA1*, *PTCHD1-AS* and *TXLNG* (Figure 2, Table 2).

**Figure 3.**
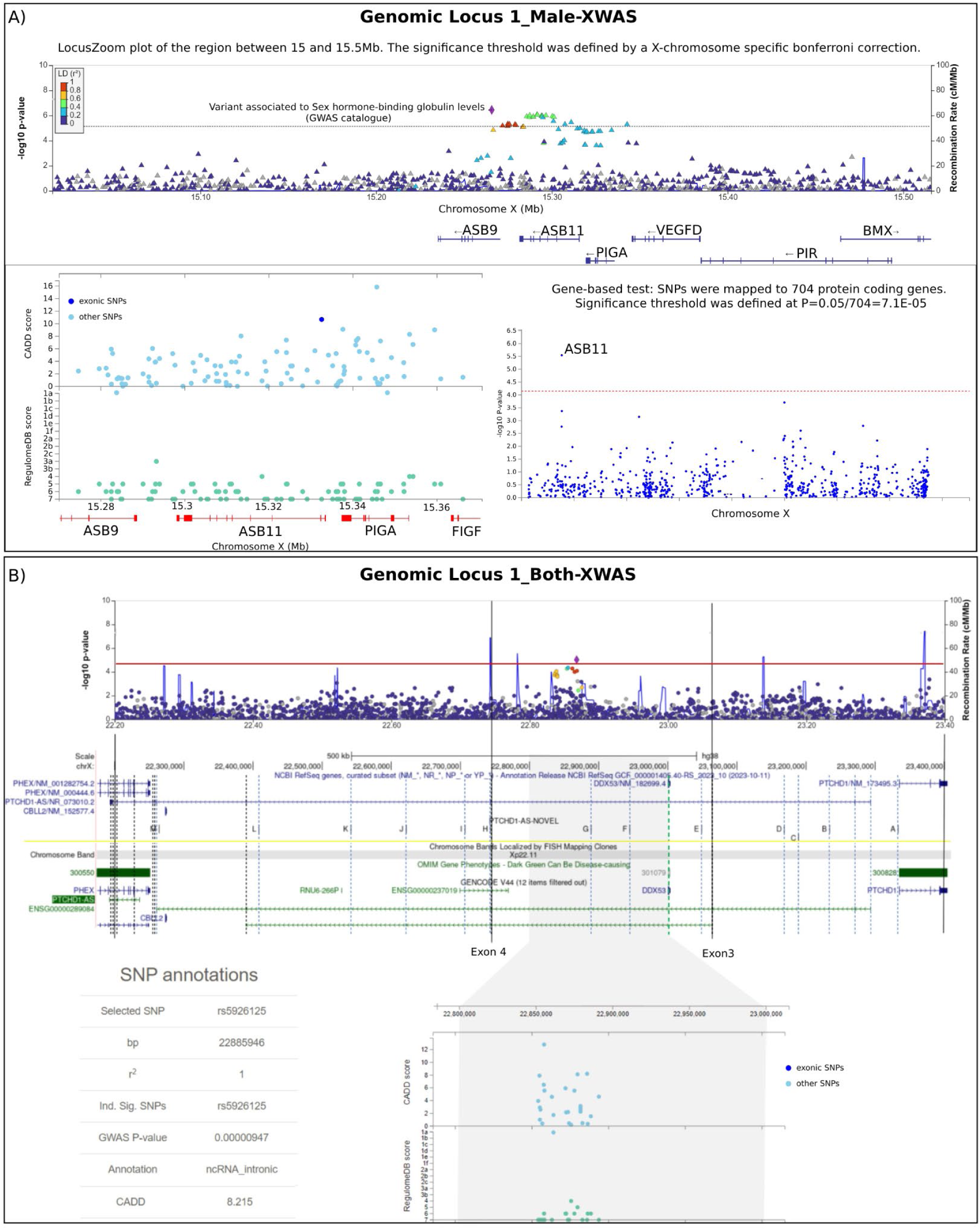
Annotation details for the genomic risk Locus 1_Male-XWAS and 1_Both XWAS. A) Details for the genomic risk locus *1_Male-XWAS*. The upper panel shows the LocusZoom plot for the correspondent region with the lead SNP rs12687599 highlighted in purple. The used LD reference panel was Europeans from 1000G data^77^ for both sexes together. Following the LocusZoom plot, on the left, we provide annotation results displaying CADD and RegulomeDB scores. On the right, the Manhattan plot illustrates the gene-based test computed by MAGMA in FUMA. The SNPs were mapped to 704 protein-coding genes, hence the genome-wide significance threshold (indicated by the red dashed line in the plot) was conservatively set at P = 0.05/704 = 7.10×10^-5^. B) LocusZoom plot of the genomic locus *1_Both-XWAS*, followed by the CADD and Regulome profiles of the same region.

In the sex-stratified analysis, the majority of the SNPs found to have significant association were located in intronic regions, accounting for 68.4% of the Male-XWAS results and 60% of the Female-XWAS results. Within the Both-XWAS results, 45.8% of the SNPs were intergenic, 28.9% were non-coding RNAs, 22.9% were intronic, and an additional 2.4% located in UTR regions.

### 3.2 Sex differences in minor allele frequencies (sdMAF)

Evolutionary forces can influence allele frequency on the X chromosome between sexes compared to the autosomes ^78,79^. To ensure the effectiveness of the quality control process, we have implemented sdMAF^60,61^ analysis on the same set of genomic data. Subsequently, we removed all sdMAF significant results from the XWAS findings. These signals could be capturing either true biological sex differences or genotyping error, inducing spurious association between ASD and variants.

However, the sdMAF results also provided valuable insights. We applied sdMAF separately to ASD individuals and controls cohorts; and we observed scatters of statistically significant variants in both ASD individuals and controls (Figure 4). Single or few scatters were expected to be caused by genotyping error. The results from the region of *FGF13* gene was particularly prominent. Notably, *FGF13* is a previously ASD-associated gene with a SFARI score of 3S (Figure 4). Interestingly, the detection of this region is solely through sdMAF but not via logistic regressions, highlighting the potential of sdMAF being used as a tool for association studies of sex-biased diagnoses.

**Figure 4.**
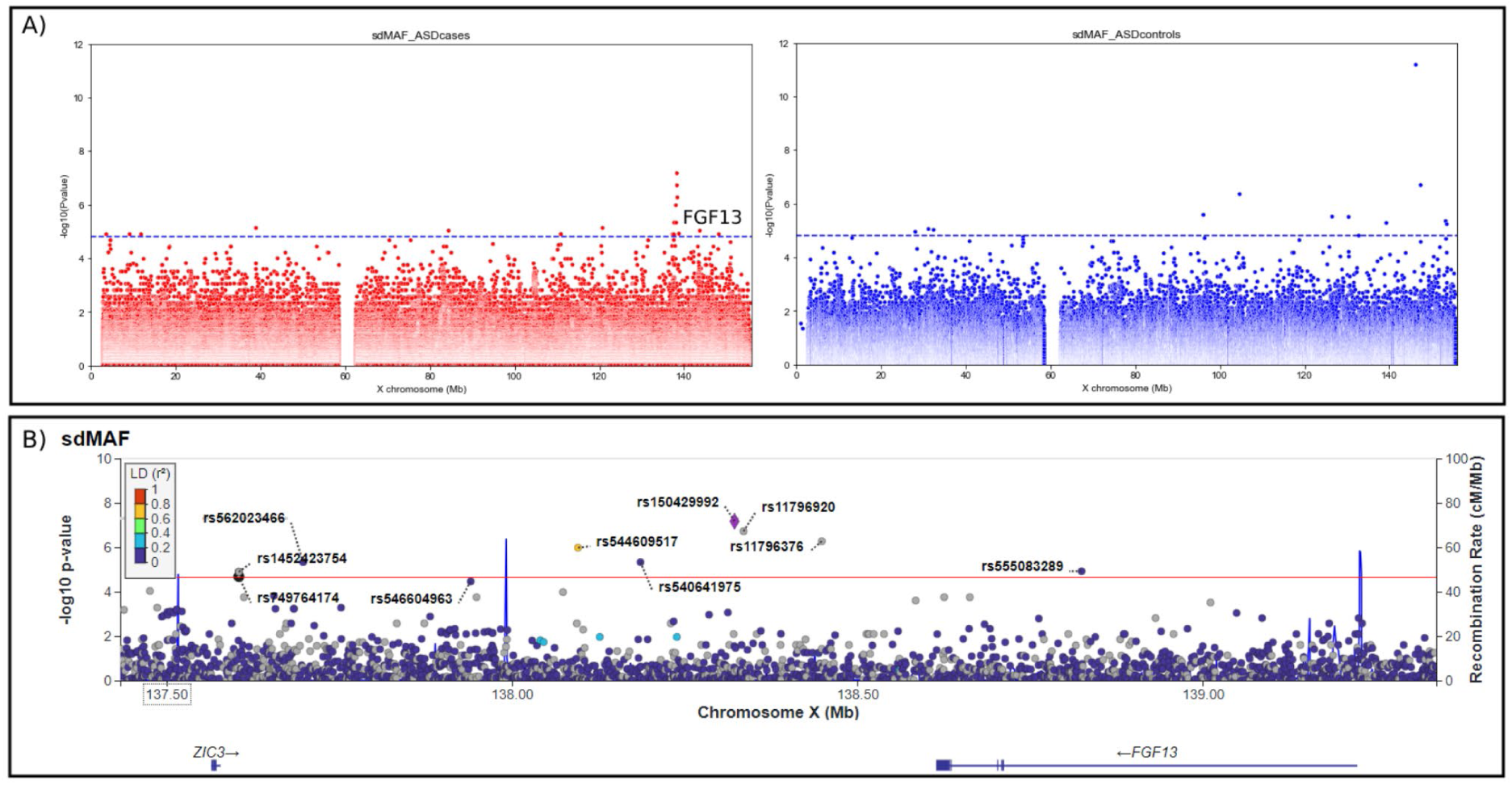
sdMaf Results. A) Left, the Manhattan plot illustrates the sdMAF p-values obtained from ASD datasets exclusively. Right, the Manhattan plot represents the sdMAF p-values obtained from control datasets only. B) The LocusZoom plot displays the region identified in the sdMAF-cases results, highlighting the gene *FGF13*. The LD reference panel used was Europeans from 1000G data^77^ for both sexes together.

### 3.3 Rare variants analysis

Recognizing the significant role of rare variants in ASD genetic architecture ^34,80–83^, we checked in the same ASD datasets (MSSNG, SSC and SPARK) for rare predicted damaging small variants (SNV/indels with less than 0.1% of frequency on gnomAD^48^) and CNV deletions (<1% frequency in gnomAD^48^) overlapping at least one exon of the 14 significant detected genes (13 from XWAS and one from sdMAF).

Among the total of 14 XWAS genes analyzed (Figure 5), 11 exhibited rare predicted damaging SNVs. Among the remaining three genes, two were non-coding RNAs (LOC124905257 at HG38 chrX:20606477:20727481 and *PTCHD1-AS* at HG38 chrX:22193005:23293146), while the third was *PABPC1L2A* (HG38 chrX: 73077276:73079512). In the male frequency comparisons, almost all genes showed a higher frequency of these variants in ASD-probands compared to other family members, except for *PCDH11X* and *PCDH19*. In females, five genes (*ASB11*, *DMD*, *HDAC8*, *PCDH19*, and *HTR2C*) showed a higher frequency in ASD-probands. Combining both sexes, four genes (*ASB11*, *DMD*, *HDAC8*, *HTR2C*, and *FGF13*) showed a higher frequency in ASD-probands.

**Figure 5.**
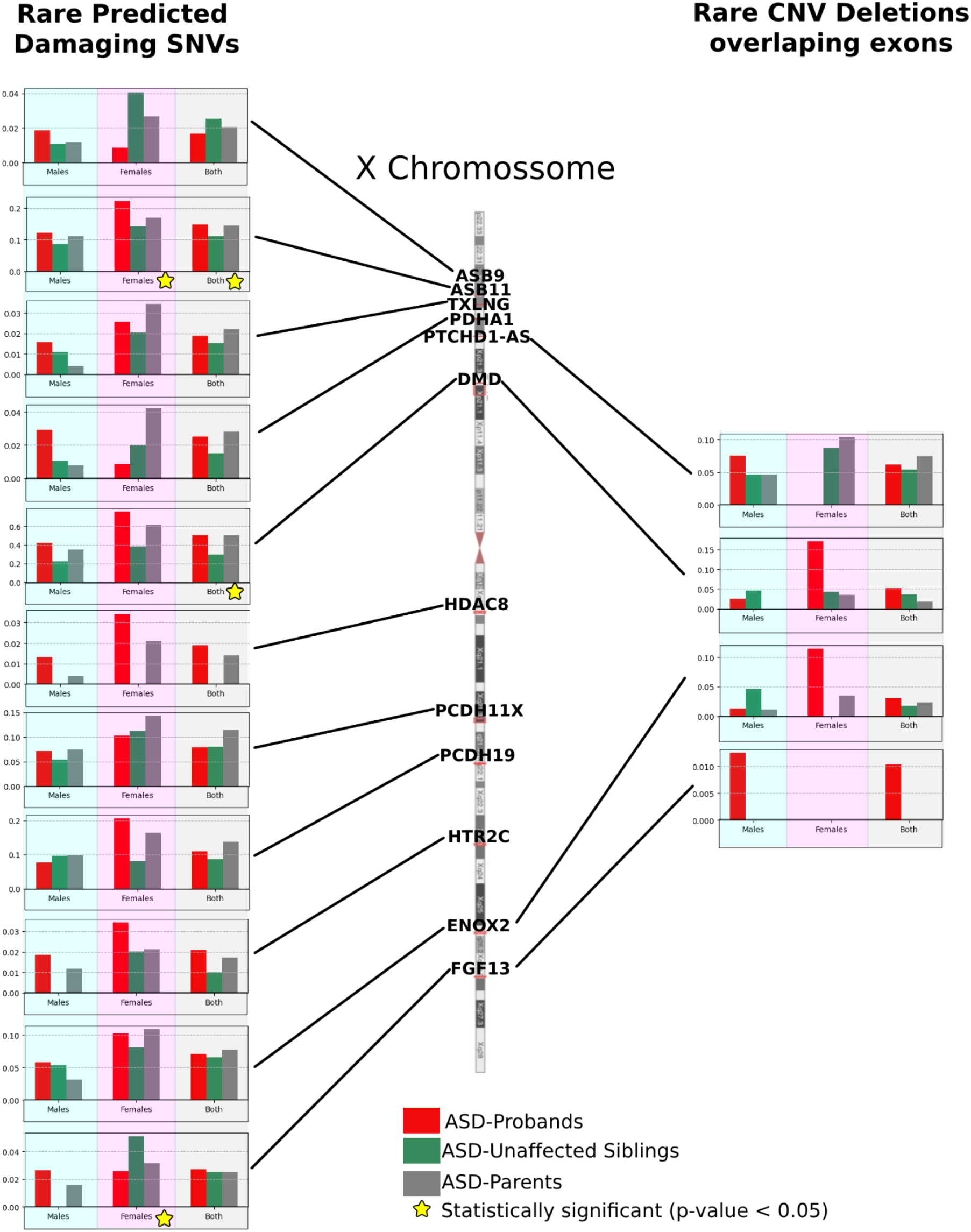
Rare Variant Frequency Analysis. The figure compares the frequencies of rare variants among different groups: ASD-Probands (red bars), ASD-Unaffected Siblings (green bars), and ASD-Parents (gray bars). The left panel shows the frequency of rare predicted damaging SNVs (<0.1% frequency in general population) across 11 genes (*ASB9*, *ASB11*, *TXLNG*, *PDHA1*, *PTCHD1-AS*, *DMD*, *HDAC8*, *PCDH11X*, *PCDH19*, *HTR2C*, *ENOX2*, *FGF13*) detected through XWAS common variant data analysis (Table 2). The right panel illustrates the frequency of rare CNV deletions overlapping exons (< 1% frequency in general population), found in four XWAS-genes (*PTCHD1-AS*, *DMD*, *ENOX2*, *FGF13*). In each graph, the corresponding p-value from a conditional logistic regression is shown at the bottom, conducted separately for males, females, and both sexes combined (using “sex” as covariate).

We successfully identified rare deletions overlapping exons in the joint ASD datasets for the gene detected in sdMAF (*FGF13*) and for three of the 13 genes from the main XWAS results, including *PTCHD1-AS, DMD*, and *ENOX2* (Figure 5, Table 2). Comparing the frequency of these CNVs in unaffected family members, we observed an enrichment in cases compared to unaffected family members for deletions impacting *PTCHD1-AS* in males, *DMD* and *ENOX2* in females and both sexes combined, and *FGF13* in males and both sexes combined. However, none of the association test results reached a p-value lower than 0.05, but this might be expected because of sample size.

### 3.4 Brain Gene expression analysis

Utilizing data from BrainSpan (Allen Brain Atlas)^71^, we generated a visualization to examine the mean expression patterns of 12 of 14 candidate genes detected in our previous analysis across various brain regions during distinct developmental periods (Figure 6). Data for *LOC124905257* and *PTCHD1-AS* were not available in BrainSpan. In general, the ASD-XWAS candidate genes showed different expression levels in all different time ranges when compared with the plotted controls (Figure 6 last three columns).

**Figure 6.**
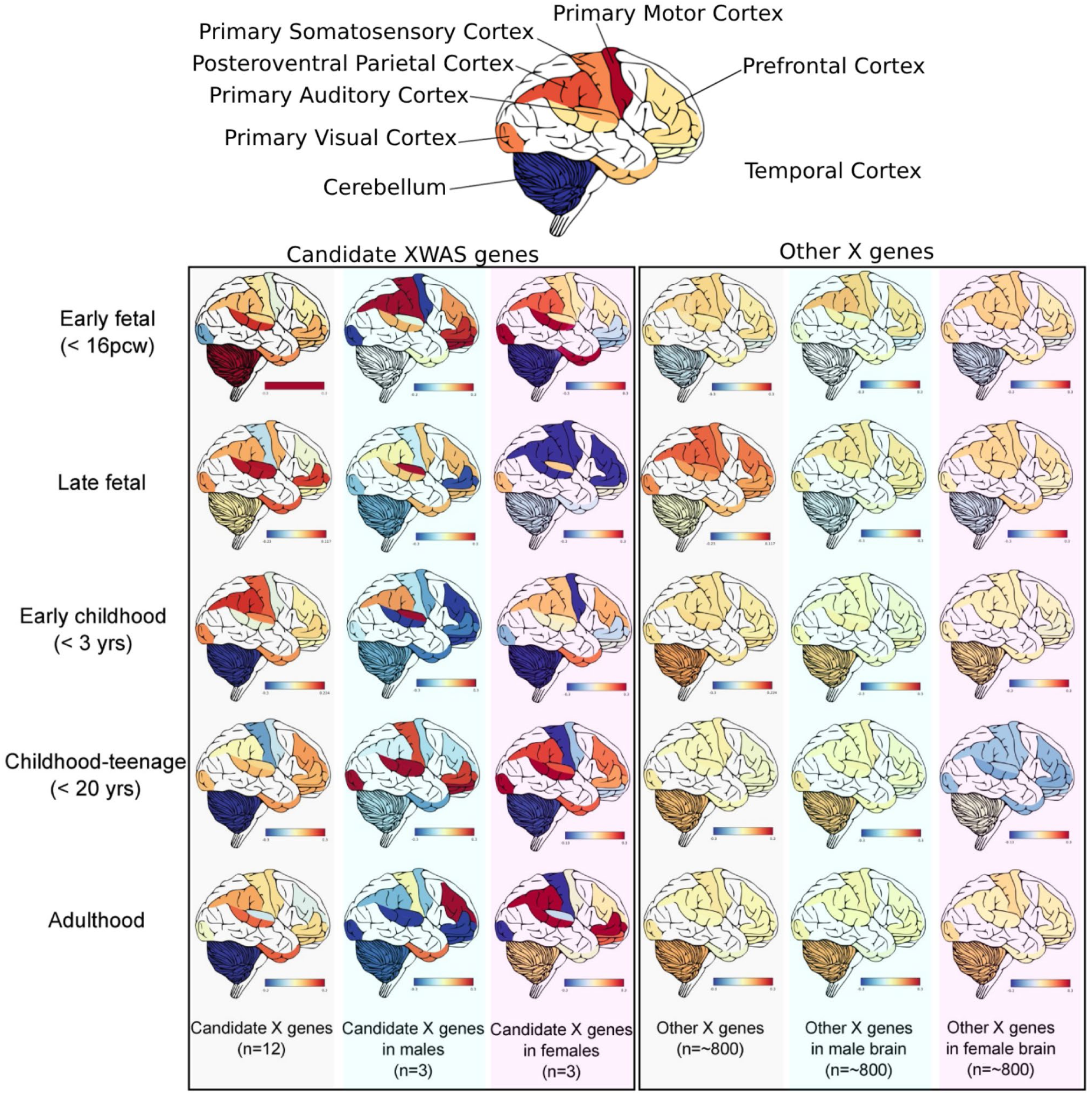
Gene Expression by Brain Regions in different development times. Brain map showing the gene expression levels in different parts of the brain in five developmental stages (Early Fetal, Late fetal, Early childhood, Childhood/Teenage and Adulthood). Left to right shows the gene expression levels from all 12 ASD-candidate genes with available expression data (*ASB11, ASB9, DMD, ENOX2, FGF13, HDAC8, HTR2C, PABPC1L2A, PCDH11X, PCDH19, PDHA1, TXLNG*), followed by three genes from Male-XWAS (*ASB11, ASB9, PCDH19*), three genes from Female-XWAS (*TXLNG*, *HTR2C*, *ENOX2*) and the correspondent control comparison with all the ∼800 X chromosome genes in both sexes and also in male brains only and female brains only. The color scales go from blue (downregulated) to red (upregulated).

During the early fetal stage, the 12 XWAS genes exhibit up-regulation in the cerebellum, which contrasts with the pattern observed in the Female-XWAS genes, showing notably low expression levels in the same region. In males, XWAS genes in the early fetal stage demonstrate down-regulated expression in the primary motor cortex and the primary visual cortex, alongside up-regulated expression in the prefrontal, primary somatosensory, and posteroventral parietal cortex. In this stage, the most expressed brain regions in Female-XWAS genes include the primary visual, primary auditory, and temporal cortex.

Transitioning to the late fetal stage, the most pronounced pattern includes down-regulated expression of Male-XWAS genes across nearly all analyzed brain regions. In contrast, All-XWAS genes exhibit heightened expression in the primary auditory, temporal, and prefrontal cortex. In early childhood, spanning the initial three years of life, a consistent down-regulated expression pattern is observed in the cerebellum across all approaches (All XWAS genes and sex-stratified comparisons). Furthermore, during this phase, the posteroventral parietal cortex displays elevated expression levels for All-XWAS genes.

From ages three to 20 (childhood to teenage years), X candidate genes remain downregulated in the cerebellum, while both sex-stratified approaches indicate up-regulation in the primary auditory and visual cortex. Additionally, the prefrontal cortex exhibits high expression levels for Male-XWAS genes.

In adulthood (after 20 years), the cerebellum maintains a down regulated pattern for all XWAS genes and for the genes identified in Male-XWAS, while exhibiting slightly higher expression levels in the genes identified in Female-XWAS. Conversely, the prefrontal cortex demonstrates low expression levels for the genes identified in Male-XWAS, with an upregulation pattern observed in the genes identified in Female-XWAS.

## 4. Discussion

Our XWAS analyses identified 59 SNP variants on the X chromosome that exhibited a statistically significant association with ASD (Table S2). These variants were mapped to 91 distinct genes, of which 11 had previously been associated with ASD through the detection of rare variants or CNVs, as reported in databases (Table S5). Out of the 59 significant variants identified in the main analysis, 35 were also successfully detected in our robustness study (Table S3), spanning all five different tests. Among these, 33 variants passed the sanity test by not reaching a significant value in the Control vs Control test “vi”. These 33 X-Chromosome variants were located in intergenic regions as well as in the genes *ASB9*, *ASB11*, *PDHA1*, *LOC124905257*, *PTCHD1*-*AS*, *HDAC8*, *PABPC1L2A*, and *PCDH11X* (Table 2).These new results will increase our understanding of the genes involved in ASD and provide a basis for improving polygenic risk scores (PRS), which currently are significantly underpowered regarding ASD^34,84^.

In the Male-XWAS results we detected an LD region encompassing the genes *ASB9*, *ASB11* and *PIGA*. The lead SNP, rs12687599, is reported in the GWAS catalog^85^ for being associated with sex hormone-binding globulin levels^86^. Autism was previously associated with a decreased level of maternal serum sex hormone binding globulin^87^. This finding could imply an etiological association between sex hormone pathways and ASD status particularly in males ^16,23^. Still in the Male-XWAS, we identified the gene *PCDH19*, also found in the Meta-XWAS. This gene has the highest significance score of 1 in the SFARI database, indicating its significant relevance to ASD^26^. It is also classified as syndromic, primarily expressed in brain tissue and plays a role in cell adhesion^88^, suggesting that mutations within it are associated with an increase in ASD likelihood and are consistently linked to neurodevelopmental and neuropsychiatric characteristics beyond those necessary for an ASD diagnosis.

The Both-XWAS and Meta-XWAS identified significantly associated variants in the lncRNA *PTCHD1-AS* (*PTCHD1* antisense RNA)^89^. This gene is part of a complex on chromosome Xp22.11, which also encompasses *DDX53*, placing this locus among the most prevalent and impactful genetic factors for ASD^90^ and other neurodevelopmental disorders. Ross *et al*., 2021^89^, conducted an analysis compiling data from previously reported variants on *PTCHD1-AS*. They found that 69% of these variants associated with this long non-coding RNA (lncRNA) are linked to ASD or ASD-related features. Consequently, the EAGLE score, a metric evaluating a gene’s relevance to ASD, definitively assigns *PTCHD1-AS* a final score of 17.6^27^. However, despite this association, the functional significance of these variants remains unknown.

In the Meta-XWAS we identified significant variants associated with ASD in *DMD* and *HDAC8*. Notably, *HDAC8* was also highlighted in the Both-XWAS results. Both genes carry a syndromic status on the SFARI gene score. Both *DMD* and *HDAC8* are linked to intellectual disability, with *DMD* additionally implicated in attention-deficit hyperactivity disorder (ADHD) and extra-pyramidal syndrome (EPS). The *DMD* gene was the only gene to reach a significant enrichment p-value (0.01) when comparing rare deletions in probands against unaffected family members specifically for females. This finding suggests a potential sex-specific effect of rare deletions in the DMD gene, with females exhibiting a significant enrichment compared to unaffected family members. Our previous genomic studies of CNVs ^34^ further support the importance of rare deletions in *DMD*.

We also applied a case-only sdMAF analysis in a complementary way to the traditional case-control association analysis. This analysis pointed out a significant peak overlapping *FGF13*^91^ with variants in this gene being involved in infantile-onset developmental and epileptic encephalopathy, which can be important associated features of ASD.

In summary, our XWAS study of individuals with ASD and controls has generated significant new data that further validate the roles of specific genes in autism and unveil novel candidates for future research. Our approach, utilizing XWAS ’common variant’ analyses alongside parallel ’rare variant’ examinations of the same samples, provides a unique paradigm for dissecting the genomic architecture involved in ASD and potentially other complex conditions. Additionally, while the development of an X-chromosome-based Polygenic Risk Score (X-PRS) is of interest, it is beyond the scope of this paper and may require new methodologies^29^.

## Data Availability

All data produced are available online at
https://research.mss.ng/
https://base.sfari.org
https://www.internationalgenome.org
https://www.cgen.ca/hostseq-databank-access-request
MGRB genomic data is deposited at the European Genome-Phenome Archive under study ID EGAS00001003511

https://research.mss.ng/

https://base.sfari.org

https://www.internationalgenome.org

https://www.cgen.ca/hostseq-databank-access-request

## Acknowledgments

We thank the families participating in MSSNG, SSC, and SPARK, as well as the resources provided by Autism Speaks and The Centre for Applied Genomics. M.M.A was supported by the CGEn HostSeq/CIHR fellowship (CGE 185054) and the SickKids Restracomp Fellowship. S.B.C was supported in part by the Intramural Research Program of the NIH and the National Institute on Aging (NIA). M.C.L is supported by a Canadian Institutes of Health Research Sex and Gender Science Chair (GSB 171373). S.W.S holds the Northbridge Chair in Pediatric Research at The Hospital for Sick Children and the University of Toronto.

## Web resources

Approved researchers can obtain the MSSNG dataset by applying at https://research.mss.ng/; and the SSC and SPARK datasets at https://base.sfari.org. 1000 genomes data is publicly available at https://www.internationalgenome.org/, HostSeq data can be also available after applying at https://www.cgen.ca/hostseq-databank-access-request and MGRB genomic data is deposited at the European Genome-Phenome Archive under study ID EGAS00001003511.

## Author contributions

Conceptualization: M.M.A, D.Z.C, T.P.L, L.S, S.W.S

Data curation: M.M.A, W.E, B.Thiruvahindrapuram, B.Trost, J.L.H, G.P, T.N

Formal Analysis: M.M.A, D.Z.C, W.E, E.A.M, S.B.C, D.R

Funding acquisition: S.W.S

Investigation: M.M.A, D.Z.C, W.E, B.Trost,, J.L.H, M.C.L, C.A.B, L.S, S.W.S

Methodology: M.M.A, D.Z.C, W.E, T.P.L, B.Thiruvahindrapuram, G.P, T.N, R.A, N.B.S, E.A.M, N.R.A, S.B.C, D.R

Project administration: J.L.H, L.S, S.W.S

Resources: S.W.S

Software: M.M.A, D.Z.C, T.P.L, L.S

Supervision: L.S, S.W.S

Validation: M.M.A, W.E, N.B.S, C.A.B

Visualization: M.M.A, W.E

Writing – original draft: M.M.A, W.E, B.Trost, M.C.L, L.S, S.W.S

Writing – review & editing: M.M.A, D.Z.C, W.E, T.P.L, B.Thiruvahindrapuram, B.Trost, J.L.H, G.P, T.N, R.A, N.B.S, E.A.M, N.R.A, M.C.L, S.B.C, D.R, C.A.B, L.S, S.W.S

