## Supplementary Materials for "Chromosome X-Wide Common Variant Association Study (XWAS) in Autism Spectrum Disorder"

**Table S1.** Comparison between our study with X chromosome and previous works that have used similar approaches for common variants.

| | | Henningsson et al., 2009 | Wang et al., 2009 | Chung et al., 2011 | Ramos et al., 2012 | Our study |
|---|---|---|---|---|---|---|
| PUBMED | | 19167832 | 19404256 | 22050706 | 22681640 | |
| Methods | Target trait | Autism | Autism | Autism | Autism | Autism |
| | "Case" Samples | 267 | 3,101 subjects and a second cohort of 1,204 affected subjects | 894 ASD families (3,128 individuals) + 939 ASD families (4,495 individuals) + 1,204 cases | 1,510 trios | 6,873 |
| | "Control" Samples | 617 | 6,491 | 6,472 | Family based test | 8,981 |
| | Sample's global ancestry | 204 from the Paris Autism Research International Sib-pair (PARIS) study and 63 came a separate Swedish cohort. | European ancestry | Self-reported Caucasian ancestry. | European descent (76%),with Asian (3%) and African (2%) | European (samples with more than 75% of european ancestry) |
| | Genetic Markers | Genotyped for three polymorphisms in exon 1 of the AR gene | 486,864 markers in all genome (not only on X chromosome) | Genotyped data (~13,837 X-chromosome SNPs) | 22,904 single nucleotide polymorphisms (SNPs) from the 2,012 immune-related genes | 418,876 variants only in the X-chromosome |
| | Association Test Method | Case-Control Comparison plus Transmition Desequilibrium test | Genome-wide association, plus family-based association test using X-APL. | Meta-analysis and joint analysis. genome-wide association study (GWAS) data on the X chromosome in three independent autism GWAS data sets. | Family-based tests of association. | XWAS |
| Results | | Case–control comparisons revealed higher prevalence of short CAG alleles as well as of the A allele of the rs6152 SNP in female cases than in controls, but revealed no significant differences with respect to the GGN repeat. | *RHOXF1* (combined pvalue 9.7×10−7), Between *PRKX* and *NLGN4X* (combined pvalue 9.0×10−7), *DMD* (combined pvalue 2.7×10−7). | One SNP, rs17321050, in the transducin b-like 1X-linked (*TBL1X*) gene showed chromosome-wide significance in the meta-analysis (P value = 4.86 × 10-6) and joint analysis (P value = 4.53 × 10-6) in males. | Significant associations in the CD99 molecule-like 2 region (CD99L2,rs11796490, P= 4.01 × 10-06, OR = 0.68 (0.58-0.80)) | Detected 59 candidate variants in 13 genes. |
| Conclusion | | The results lend some support for an influence of the studied polymorphisms on the susceptibility for autism, but argue against the possibility that mutations in the AR gene are common in subjects with this condition. | The first demonstration of genome-wide significant association of common variants with susceptibility to ASDs including X chromosome. | Our results, based on meta-analysis, joint analysis and replication analysis, suggest that *TBL1X* may play a role in ASD risk. | Several novel genes with known immune functions are associated with ASD. | First XWAS using WGS data in Autism, finding variants in 6 ASD-genes plus 8 new ASD-genes candidates. |

**Table S2.** Significant associated variants with the respective P-Value from all different XWAS (XWAS-Males, XWAS-Females, XWAS-Both, XWAS-Meta).

**Table S3.** Significant associated variants with the respective P-Value from all different XWAS (XWAS-Males, XWAS-Females, XWAS-Both, XWAS-Meta) from all replication tests: i. MSSNG as case vs HostSeq and MGRB as controls; ii. SSC as case vs HostSeq and MGRB as controls; iii. SPARK as case vs HostSeq and MGRB as controls; iv. MSSNG, SSC and SPARK as cases vs HostSeq as controls; v. MSSNG, SSC and SPARK as case and MGRB as controls.

**Table S4.** Significant associated variants with the respective P-Value from all different XWAS (Male-XWAS, female-XWAS, Both-XWAS and Meta-XWAS) when using the 10 top autosomal principal components as covariates.

**Table S5.** The 91 mapped genes (10kb distance) based on the XWAS significant variants. In red are highlighted the genes previously associated with ASD on both SFARI and Autdb, and in yellow the genes that was presented in at least one of those 2 autism gene score systems.

| Gene | Count | IndSigSNPs | Males | Females | Both | Meta Analys | SFARI Score | Autdb gene |
|---|---|---|---|---|---|---|---|---|
| GRPR | 3 | rs6628945 | 8.27E-07 | 6.01E-01 | 1.12E-03 | 1.22E-03 | | 2 |
| RAI2 | 3 | rs12687599 | 3.57E-07 | 6.79E-01 | 5.56E-04 | 5.74E-04 | | |
| ZRSR2 | 3 | rs6628945 | 8.27E-07 | 6.01E-01 | 1.12E-03 | 1.22E-03 | | |
| AIFM1 | 2 | rs186814690 | 2.68E-02 | 2.41E-06 | 5.59E-05 | 5.06E-07 | | |
| AP1S2 | 2 | rs185190849 | 5.08E-01 | 1.68E-06 | 9.01E-03 | 1.31E-03 | Syndromic | 2 |
| APLN | 2 | rs186814690 | 2.68E-02 | 2.41E-06 | 5.59E-05 | 5.06E-07 | | |
| ARHGAP36 | 2 | rs186814690 | 2.68E-02 | 2.41E-06 | 5.59E-05 | 5.06E-07 | | |
| ASB11 | 2 | rs12687599; rs6628945 | 3.57E-07 | 6.79E-01 | 5.56E-04 | 5.74E-04 | | |
| CA5B | 2 | rs6628945 | 8.27E-07 | 6.01E-01 | 1.12E-03 | 1.22E-03 | | |
| DDX53 | 2 | rs5926125 | 1.89E-04 | 1.04E-02 | 9.47E-06 | 7.30E-06 | 2 | 2 |
| ENOX2 | 2 | rs186814690 | 2.68E-02 | 2.41E-06 | 5.59E-05 | 5.06E-07 | | |
| GPR119 | 2 | rs186814690 | 2.68E-02 | 2.41E-06 | 5.59E-05 | 5.06E-07 | | |
| HDAC8 | 2 | rs5958792 | 1.38E-04 | 2.57E-04 | 1.85E-06 | 1.73E-07 | Syndromic | 2 |
| HTR2C | 2 | rs191071511 | 1.08E-01 | 9.58E-06 | 7.07E-03 | 3.26E-06 | | |
| MBTPS2 | 2 | rs767542284 | 7.93E-04 | 2.62E-03 | 2.61E-06 | 3.01E-05 | | |
| OR13H1 | 2 | rs186814690 | 2.68E-02 | 2.41E-06 | 5.59E-05 | 5.06E-07 | | |
| PCDH19 | 2 | rs12835197; rs5926125 | 4.33E-06 | 8.30E-02 | 3.94E-04 | 4.87E-06 | 1 | 4 |
| PHEX | 2 | rs5926125; rs189525731; rs186125715 | 1.89E-04 | 1.04E-02 | 9.47E-06 | 7.30E-06 | | |
| PIGA | 2 | rs6628945; rs12687599; rs185190849 | 8.27E-07 | 6.01E-01 | 1.12E-03 | 1.22E-03 | | |
| PTCHD1 | 2 | rs5926125; rs5945876 | 1.89E-04 | 1.04E-02 | 9.47E-06 | 7.30E-06 | 1 | 3 |
| PCDH11X | 1 | rs184604300 | 2.25E-04 | 2.12E-02 | 4.92E-03 | 1.38E-05 | 2 | 2 |
| PTCHD1-AS | 2 | rs5926125; rs5945876 | 1.89E-04 | 1.04E-02 | 9.47E-06 | 7.30E-06 | 2 | |
| RAB33A | 2 | rs186814690; rs111827716 | 2.68E-02 | 2.41E-06 | 5.59E-05 | 5.06E-07 | | |
| RBMX2 | 2 | rs186814690 | 2.68E-02 | 2.41E-06 | 5.59E-05 | 5.06E-07 | | |
| S100G | 2 | rs185190849 | 1.31E-03 | 5.08E-01 | 1.68E-06 | 9.01E-03 | | |
| SCML1 | 2 | rs773693168 | 7.96E-01 | 9.80E-06 | 3.75E-03 | 1.63E-03 | | |
| SRPX2 | 2 | rs12835197; rs186125715: rs189525731 | 4.33E-06 | 8.30E-02 | 3.94E-04 | 4.87E-06 | | |
| TMEM27 | 2 | rs6628945; rs185190849 | 8.27E-07 | 6.01E-01 | 1.12E-03 | 1.22E-03 | | |
| TNMD | 2 | rs12835197; rs189525731 | 4.33E-06 | 8.30E-02 | 3.94E-04 | 4.87E-06 | | |
| TSPAN6 | 2 | rs12835197 | 4.33E-06 | 8.30E-02 | 3.94E-04 | 4.87E-06 | | |
| YY2 | 2 | rs767542284; rs776360992 | 7.93E-04 | 2.62E-03 | 2.61E-06 | 3.01E-05 | | |
| ZNF280C | 2 | rs186814690; rs5945876 | 2.68E-02 | 2.41E-06 | 5.59E-05 | 5.06E-07 | | |
| ZNF645 | 2 | rs5926125; rs189525731 | 7.30E-06 | 1.89E-04 | 1.04E-02 | 9.47E-06 | | |
| DMD | 1 | rs5945876 | 1.23E-02 | 1.26E-04 | 1.84E-05 | 9.79E-06 | Syndromic | 3 |
| TRAPPC2 | 1 | rs6628945:rs12687599 | 8.27E-07 | 6.01E-01 | 1.12E-03 | 1.22E-03 | | |
| SYAP1 | 1 | rs185190849 | 1.31E-03 | 5.08E-01 | 1.68E-06 | 9.01E-03 | 2 | yes |
| CNKSR2 | 1 | rs5945876 | 1.23E-02 | 1.26E-04 | 1.84E-05 | 9.79E-06 | 2 | 4 |
| GLRA2 | 1 | rs12687599 | 3.57E-07 | 6.79E-01 | 5.56E-04 | 5.74E-04 | 2 | 3 |
| OFD1 | 1 | rs6628945:rs12687599 | 8.27E-07 | 6.01E-01 | 1.12E-03 | 1.22E-03 | 2 | 3 |
| CDKL5 | 1 | rs767542284 | 7.93E-04 | 2.62E-03 | 2.61E-06 | 3.01E-05 | 1 | 4 |
| GPRASP2 | 1 | rs140894960 | 7.01E-02 | 9.58E-06 | 3.95E-03 | 1.76E-06 | | yes |
| NXF5 | 1 | rs5945876 | 1.23E-02 | 1.26E-04 | 1.84E-05 | 9.79E-06 | | yes |
| SH3KBP1 | 1 | rs767542284 | 7.93E-04 | 2.62E-03 | 2.61E-06 | 3.01E-05 | | 2 |
| ACE2 | 1 | rs6628945:rs12687599 | 8.27E-07 | 6.01E-01 | 1.12E-03 | 1.22E-03 | | |
| ARMCX1 | 1 | rs189525731; rs186125715; rs186125715: rs189525731 | 1.63E-02 | 3.32E-06 | 3.86E-05 | 3.40E-07 | | |
| ARMCX2 | 1 | rs5926125 | 1.89E-04 | 1.04E-02 | 9.47E-06 | 7.30E-06 | | |
| ARMCX3 | 1 | rs139802025 | 8.35E-05 | 7.82E-03 | 5.33E-04 | 2.44E-06 | | |

| Gene | Count | IndSigSNPs | Males | Females | Both | Meta Analys |
|---|---|---|---|---|---|---|
| ARMCX5 | 1 | rs5945876 | 1.23E-02 | 1.26E-04 | 1.84E-05 | 9.79E-06 |
| ARMCX6 | 1 | rs5945876 | 1.23E-02 | 1.26E-04 | 1.84E-05 | 9.79E-06 |
| ASB9 | 1 | rs12687599; rs12687599: rs6628945 | 3.57E-07 | 6.79E-01 | 5.56E-04 | 5.74E-04 |
| BEX5 | 1 | rs776360992 | 2.60E-05 | 5.11E-02 | 4.29E-05 | 6.15E-06 |
| BHLHB9 | 1 | rs73218354 | 8.14E-05 | 3.92E-04 | 1.04E-05 | 1.42E-07 |
| BMX | 1 | rs6628945:rs12687599 | 8.27E-07 | 6.01E-01 | 1.12E-03 | 1.22E-03 |
| CTPS2 | 1 | rs185190849 | 5.08E-01 | 1.68E-06 | 9.01E-03 | 1.31E-03 |
| CXorf23 | 1 | rs767542284 | 7.93E-04 | 2.62E-03 | 2.61E-06 | 3.01E-05 |
| EGFL6 | 1 | rs12687599:rs6628945 | 3.57E-07 | 6.79E-01 | 5.56E-04 | 5.74E-04 |
| EIF1AX | 1 | rs767542284 | 7.93E-04 | 2.62E-03 | 2.61E-06 | 3.01E-05 |
| ELF4 | 1 | rs73218354;rs5958792 | 8.14E-05 | 3.92E-04 | 1.04E-05 | 1.42E-07 |
| FANCB | 1 | rs12687599 | 3.57E-07 | 6.79E-01 | 5.56E-04 | 5.74E-04 |
| FIGF | 1 | rs6628945 | 8.27E-07 | 6.01E-01 | 1.12E-03 | 1.22E-03 |
| GEMIN8 | 1 | rs12687599:rs6628945 | 3.57E-07 | 6.79E-01 | 5.56E-04 | 5.74E-04 |
| GPM6B | 1 | rs12687599:rs6628945 | 3.57E-07 | 6.79E-01 | 5.56E-04 | 5.74E-04 |
| GPRASP1 | 1 | rs5926125 | 1.89E-04 | 1.04E-02 | 9.47E-06 | 7.30E-06 |
| IGSF1 | 1 | rs73218354 | 8.14E-05 | 3.92E-04 | 1.04E-05 | 1.42E-07 |
| KLHL34 | 1 | rs111827716 | 5.63E-04 | 4.36E-03 | 4.45E-04 | 8.40E-06 |
| MAGEB17 | 1 | rs185190849 | 5.08E-01 | 1.68E-06 | 9.01E-03 | 1.31E-03 |
| MAP3K15 | 1 | rs767542284 | 7.93E-04 | 2.62E-03 | 2.61E-06 | 3.01E-05 |
| MAP7D2 | 1 | rs767542284 | 7.93E-04 | 2.62E-03 | 2.61E-06 | 3.01E-05 |
| MOSPD2 | 1 | rs12687599 | 3.57E-07 | 6.79E-01 | 5.56E-04 | 5.74E-04 |
| NAP1L2 | 1 | rs776360992:rs5926125 | 2.60E-05 | 5.11E-02 | 4.29E-05 | 6.15E-06 |
| NHS | 1 | rs773693168 | 7.96E-01 | 9.80E-06 | 3.75E-03 | 1.63E-03 |
| PABPC1L2A | 1 | rs186125715:rs189525731 | 3.79E-02 | 1.04E-05 | 1.51E-04 | 2.37E-06 |
| PABPC1L2B | 1 | rs186125715; rs186125715: rs189525731 | 3.79E-02 | 1.04E-05 | 1.51E-04 | 2.37E-06 |
| PDHA1 | 1 | rs767542284 | 7.93E-04 | 2.62E-03 | 2.61E-06 | 3.01E-05 |
| PHKA2 | 1 | rs767542284 | 7.93E-04 | 2.62E-03 | 2.61E-06 | 3.01E-05 |
| PIR | 1 | rs6628945:rs12687599 | 8.27E-07 | 6.01E-01 | 1.12E-03 | 1.22E-03 |
| PLS3 | 1 | rs5945876 | 1.23E-02 | 1.26E-04 | 1.84E-05 | 9.79E-06 |
| PPEF1 | 1 | rs767542284 | 7.93E-04 | 2.62E-03 | 2.61E-06 | 3.01E-05 |
| PRPS2 | 1 | rs12687599:rs6628945 | 3.57E-07 | 6.79E-01 | 5.56E-04 | 5.74E-04 |
| RAB40AL | 1 | rs186125715 | 3.79E-02 | 1.04E-05 | 1.51E-04 | 2.37E-06 |
| RAB9A | 1 | rs12687599:rs6628945 | 3.57E-07 | 6.79E-01 | 5.56E-04 | 5.74E-04 |
| RBBP7 | 1 | rs773693168 | 7.96E-01 | 9.80E-06 | 3.75E-03 | 1.63E-03 |
| REPS2 | 1 | rs773693168 | 7.96E-01 | 9.80E-06 | 3.75E-03 | 1.63E-03 |
| RS1 | 1 | rs767542284 | 7.93E-04 | 2.62E-03 | 2.61E-06 | 3.01E-05 |
| SCML2 | 1 | rs767542284 | 7.93E-04 | 2.62E-03 | 2.61E-06 | 3.01E-05 |
| TAB3 | 1 | rs12835197 | 4.33E-06 | 8.30E-02 | 3.94E-04 | 4.87E-06 |
| TCEAL2 | 1 | rs111827716 | 5.63E-04 | 4.36E-03 | 4.45E-04 | 8.40E-06 |
| TCEAL6 | 1 | rs5945876 | 1.23E-02 | 1.26E-04 | 1.84E-05 | 9.79E-06 |
| TCEANC | 1 | rs6628945:rs12687599 | 8.27E-07 | 6.01E-01 | 1.12E-03 | 1.22E-03 |
| TIMM8A | 1 | rs186125715:rs189525731 | 3.79E-02 | 1.04E-05 | 1.51E-04 | 2.37E-06 |
| TMSB15A | 1 | rs776360992 | 2.60E-05 | 5.11E-02 | 4.29E-05 | 6.15E-06 |
| TXLNG | 1 | rs185190849 | 1.31E-03 | 5.08E-01 | 1.68E-06 | 9.01E-03 |
| ZMAT1 | 1 | rs5945876 | 1.23E-02 | 1.26E-04 | 1.84E-05 | 9.79E-06 |

**Table S6. SFARI and EAGLE genes on X previously implicated in ASD.** In red are highlighted the genes that were also found in our X-chromosome results with XWAS approach (13 genes), and sdMAF (1 gene).

| gene-symbol | gene-name | chromosome | genetic-category | gene-score | syndromic | eagle | number-of-reports |
|---|---|---|---|---|---|---|---|
| AFF2 | AF4/FMR2 family, member 2 | X | Rare Single Gene Mutation, Syndromic | 1 | 0 | 8.7 | 21 |
| AGTR2 | angiotensin II receptor, type 2 | X | Rare Single Gene Mutation | 2 | 0 | | 5 |
| AP1S2 | adaptor related protein complex 1 sigma 2 subunit | X | Rare Single Gene Mutation, Syndromic | | 1 | | 7 |
| ARHGEF9 | Cdc42 guanine nucleotide exchange factor (GEF) 9 | X | Rare Single Gene Mutation, Syndromic | 1 | 1 | 14.2 | 17 |
| ARX | aristaless related homeobox | X | Rare Single Gene Mutation, Syndromic | 1 | 1 | 13.8 | 26 |
| AR | androgen receptor | X | Genetic Association | 2 | 0 | | 6 |
| ASMT | acetylserotonin O-methyltransferase | X,Y | Rare Single Gene Mutation, Genetic Association | 2 | 0 | | 11 |
| ATRX | alpha thalassemia/mental retardation syndrome X-linked | X | Rare Single Gene Mutation, Syndromic, Functional | 1 | 0 | | 32 |
| BCORL1 | BCL6 corepressor like 1 | X | Rare Single Gene Mutation, Syndromic | | 1 | | 8 |
| BRWD3 | bromodomain and WD repeat domain containing 3 | X | Rare Single Gene Mutation, Syndromic | | 1 | | 9 |
| CACNA1F | calcium channel, voltage-dependent, alpha 1F | X | Rare Single Gene Mutation, Genetic Association | 2 | 0 | | 9 |
| CASK | calcium/calmodulin dependent serine protein kinase | X | Rare Single Gene Mutation, Syndromic, Functional | 1 | 0 | | 25 |
| CD99L2 | CD99 molecule like 2 | X | Genetic Association | 2 | 0 | | 1 |
| CDK16 | cyclin dependent kinase 16 | X | Rare Single Gene Mutation | 2 | 0 | | 3 |
| CDKL5 | cyclin-dependent kinase-like 5 | X | Rare Single Gene Mutation, Syndromic, Functional | 1 | 1 | | 55 |
| CHM | CHMRab escort protein | X | Rare Single Gene Mutation | 3 | 0 | | 7 |
| CLCN4 | chloride voltage-gated channel 4 | X | Rare Single Gene Mutation, Syndromic | 2 | 1 | | 11 |
| CNKSR2 | connector enhancer of kinase suppressor of Ras 2 | X | Rare Single Gene Mutation, Syndromic, Functional | 2 | 1 | | 12 |
| DDX3X | DEAD (Asp-Glu-Ala-Asp) box helicase 3, X-linked | X | Rare Single Gene Mutation, Syndromic, Functional | 1 | 1 | | 48 |
| DDX53 | DEAD (Asp-Glu-Ala-Asp) box polypeptide 53 | X | Rare Single Gene Mutation | 2 | 0 | 5.25 | 5 |
| DMD | dystrophin (muscular dystrophy, Duchenne and Becker types) | X | Rare Single Gene Mutation, Syndromic, Genetic Association | | 1 | | 43 |
| FAM47A | family with sequence similarity 47 member A | X | Rare Single Gene Mutation | 2 | 0 | 0.45 | 1 |
| FGF13 | fibroblast growth factor 13 | X | Syndromic | 3 | 1 | | 2 |
| FLNA | filamin A | X | Rare Single Gene Mutation, Functional | 3 | 0 | | 9 |
| FMR1 | fragile X mental retardation 1 | X | re Single Gene Mutation, Syndromic, Genetic Association, Functio | 1 | 1 | | 63 |
| FRMPD4 | FERM and PDZ domain containing 4 | X | Rare Single Gene Mutation, Syndromic | | 1 | | 8 |
| GABRA3 | Gamma-aminobutyric acid (GABA) A receptor, alpha 3 | X | Rare Single Gene Mutation, Syndromic | | 1 | | 3 |
| GLRA2 | glycine receptor, alpha 2 | X | Rare Single Gene Mutation, Functional | 2 | 0 | | 17 |
| GPC4 | glypican 4 | X | Rare Single Gene Mutation | 2 | 0 | | 2 |
| GRIA3 | glutamate ionotropic receptor AMPA type subunit 3 | X | Rare Single Gene Mutation, Syndromic, Functional | | 1 | | 15 |
| HCFC1 | host cell factor C1 | X | Rare Single Gene Mutation, Syndromic | | 1 | | 12 |
| HDAC8 | histone deacetylase 8 | X | Rare Single Gene Mutation, Syndromic | | 1 | | 11 |
| HNRNPH2 | heterogeneous nuclear ribonucleoprotein H2 | X | Rare Single Gene Mutation, Syndromic | 1 | 0 | | 19 |
| HUWE1 | CT, UBA and WWE domain containing 1, E3 ubiquitin protein liga | X | Rare Single Gene Mutation, Syndromic | | 1 | | 25 |
| IL1RAPL1 | interleukin 1 receptor accessory protein-like 1 | X | Rare Single Gene Mutation | 2 | 0 | | 27 |
| IL1RAPL2 | interleukin 1 receptor accessory protein-like 2 | X | Rare Single Gene Mutation, Genetic Association | 2 | 0 | | 2 |
| IQSEC2 | IQ motif and Sec7 domain 2 | X | Rare Single Gene Mutation, Syndromic, Functional | 1 | 1 | | 49 |
| KDM5C | lysine demethylase 5C | X | Rare Single Gene Mutation, Syndromic, Functional | 1 | 0 | | 38 |
| KDM6A | lysine demethylase 6A | X | Rare Single Gene Mutation, Syndromic | 2 | 0 | | 14 |
| LAS1L | LAS1 like ribosome biogenesis factor | X | Rare Single Gene Mutation, Syndromic | 3 | 0 | | 5 |
| MAOA | monoamine oxidase A | X | re Single Gene Mutation, Syndromic, Genetic Association, Functio | 2 | 0 | | 20 |
| MAOB | monoamine oxidase B | X | re Single Gene Mutation, Syndromic, Genetic Association, Functio | 2 | 0 | | 6 |
| MECP2 | Methyl CpG binding protein 2 | X | Rare Single Gene Mutation, Syndromic, Functional | 1 | 1 | 106.65 | 119 |
| MSL3 | MSL complex subunit 3 | X | Rare Single Gene Mutation, Syndromic | 1 | 1 | | 6 |
| NAA10 | N-alpha-acetyltransferase 10, NatA catalytic subunit | X | Rare Single Gene Mutation, Syndromic | 3 | 1 | | 13 |
| NEXMIF | neurite extension and migration factor | X | Rare Single Gene Mutation, Syndromic, Functional | 1 | 0 | | 33 |
| NLGN3 | neuroligin 3 | X | Rare Single Gene Mutation, Genetic Association, Functional | 1 | 0 | 6.5 | 45 |
| NLGN4X | neuroligin 4, X-linked | X | re Single Gene Mutation, Syndromic, Genetic Association, Functio | 1 | 0 | 12 | 42 |
| OCRL | oculocerebrorenal syndrome of Lowe | X | Rare Single Gene Mutation, Syndromic | | 1 | | 9 |
| OFD1 | OFD1, centriole and centriolar satellite protein | X | Rare Single Gene Mutation, Syndromic | 2 | 0 | | 5 |
| OPHN1 | oligophrenin 1 | X | Rare Single Gene Mutation, Syndromic | 2 | 0 | | 18 |
| PCDH11X | protocadherin 11 X-linked | X | Rare Single Gene Mutation | 2 | 0 | | 4 |
| PCDH19 | protocadherin 19 | X | Rare Single Gene Mutation, Syndromic, Functional | 1 | 1 | | 56 |
| PHF8 | PHD finger protein 8 | X | Rare Single Gene Mutation, Syndromic | | 1 | | 13 |
| PJA1 | praja ring finger ubiquitin ligase 1 | X | Syndromic | 3 | 1 | | 1 |
| PLXNA3 | plexin A3 | X | Rare Single Gene Mutation, Syndromic | 2 | 0 | | 8 |
| PTCHD1 | patched domain containing 1 | X | Rare Single Gene Mutation, Genetic Association | 1 | 0 | 0 | 15 |
| PTCHD1-AS | PTCHD1antisense RNA (head to head) | X | Rare Single Gene Mutation | 2 | 0 | 17.6 | 3 |
| RAB39B | RAB39B, member RAS oncogene family | X | Rare Single Gene Mutation, Syndromic, Functional | 2 | 0 | | 19 |
| RHOXF1 | Rhox homeobox family, member 1 | X | Genetic Association | 2 | 0 | | 3 |
| RLIM | Ring finger protein, LIM domain interacting | X | Rare Single Gene Mutation, Syndromic | | 1 | | 6 |
| RPL10 | ribosomal protein L10 | X | Rare Single Gene Mutation, Syndromic | 2 | 0 | | 14 |
| RPS6KA3 | Ribosomal protein S6 kinase, 90kDa, polypeptide 3 | X | Rare Single Gene Mutation, Syndromic | 2 | 1 | | 18 |
| SHOX | short stature homeobox | X,Y | Rare Single Gene Mutation | 2 | 0 | | 3 |
| SLC6A8 | te carrier family 6 (neurotransmitter transporter, creatine), mem | X | Rare Single Gene Mutation, Syndromic | 2 | 0 | | 24 |
| SLC7A3 | carrier family 7 (cationic amino acid transporter, y+ system), me | X | Rare Single Gene Mutation | 2 | 0 | | 2 |
| SLC9A6 | solute carrier family 9 (sodium/hydrogen exchanger), member 6 | X | Rare Single Gene Mutation, Syndromic, Functional | 1 | 1 | | 21 |
| SLITRK2 | SLIT and NTRK like family member 2 | X | Rare Single Gene Mutation, Syndromic, Functional | | 1 | | 7 |
| SMC1A | structural maintenance of chromosomes 1A | X | Rare Single Gene Mutation, Syndromic | | 1 | | 16 |
| SYAP1 | Synapse associated protein 1 | X | Rare Single Gene Mutation | 2 | 0 | | 2 |
| SYN1 | Synapsin 1 | X | Rare Single Gene Mutation, Functional | 1 | 0 | 2.35 | 27 |
| SYP | synaptophysin | X | Rare Single Gene Mutation | 3 | 0 | | 4 |
| TAF1 | TATA-box binding protein associated factor 1 | X | Rare Single Gene Mutation, Syndromic | | 1 | | 17 |
| TBX22 | T-box transcription factor 22 | X | Rare Single Gene Mutation | 3 | 0 | | 2 |
| TCEAL1 | transcription elongation factor A like 1 | X | Rare Single Gene Mutation, Syndromic | 3 | 1 | | 2 |
| TFE3 | transcription factor binding to IGHM enhancer 3 | X | Rare Single Gene Mutation, Syndromic | | 1 | | 5 |
| TMLHE | trimethyllysine hydroxylase, epsilon | X | Rare Single Gene Mutation, Genetic Association, Functional | 2 | 0 | | 10 |
| TRPC5 | ransient receptor potential cation channel subfamily C member | X | Rare Single Gene Mutation | 3 | 0 | | 7 |
| TSPAN7 | tetraspanin 7 | X | Rare Single Gene Mutation, Functional | 2 | 0 | | 9 |
| TBL1X | transducin (beta)-like 1X-linked | X | Genetic Association | 2 | 0 | | 1 |
| UPF3B | UPF3B, regulator of nonsense mediated mRNA decay | X | Rare Single Gene Mutation, Syndromic | 1 | 1 | | 17 |
| USP9X | ubiquitin specific peptidase 9 X-linked | X | Rare Single Gene Mutation, Syndromic, Functional | 1 | 1 | | 15 |
| VSIG4 | V-set and immunoglobulin domain containing 4 | X | Rare Single Gene Mutation | 2 | 0 | | 3 |
| WNK3 | WNK lysine deficient protein kinase 3 | X | Rare Single Gene Mutation | 2 | 0 | | 6 |
| ZMYM3 | zinc finger MYM-type containing 3 | X | Rare Single Gene Mutation, Syndromic, Genetic Association | | 1 | | 8 |
| ZNF711 | zinc finger protein 711 | X | Rare Single Gene Mutation | 2 | 0 | | 5 |

### Autosomes Quality Control Process

| Autosomes | | |
|---|---|---|
| After QC: Filter biallelic variants that are common with the ancestry reference population, plus SmartQC | | |
| | Samples | Variants |
| MSSNG | 9,620 | 1,828,586 |
| SSC | 9,209 | 1,847,146 |
| SPARK | 12,519 | 1,833,775 |
| MGRB | 2,572 | 1,629,895 |
| HostSeq | 10,023 | 1,726,650 |
| 1KGP | 3,202 | 1,864,496 |

⬇

| | Samples | Variants |
|---|---|---|
| Merge MSSNG, SSC, SPARK, MGRB, HostSeq | 22,242 | 1,407,803 |
| XWAS Cleaning | 21,089 | 1,075,065 |
| Ancetry Check | | |
| Merge with 1KGP | 24,291 | 1,062,930 |
| LD filter | 24,291 | 132,551 |
| Remove problematic regions | 24,291 | 131,291 |
| list samples >75% European ancestry based on ADMIXTURE results | 15,854 | |
| Filter samples >75% european ancestry based on ADMIXTURE results | 15,854 | 1,075,065 |
| XWAS final input file | 15,854 | 1,075,065 |

### X Chromosome Quality Control Process

| X chromosome | | | | |
|---|---|---|---|---|
| | Begin | | After smartQC | |
| | Samples | Variants | Samples | Variants |
| MSSNG | 9,621 | 6,539,590 | 9,552 | 3,919,536 |
| SSC | 9,209 | 4,940,291 | 9,172 | 3,356,074 |
| SPARK | 12,519 | 6,328,900 | 12,474 | 3,947,717 |
| MGRB | 2,572 | 2,645,253 | 2,561 | 1,739,780 |
| HostSeq | 10,000 | 2,389,836 | 9,802 | 1,455,047 |

⬇

| | Samples | Variants |
|---|---|---|
| Merge MSSNG, SSC, SPARK, MGRB, HostSeq | 22,242 | 538,856 |
| XWAS Cleaning | 21,089 | 418,652 |
| Filter samples >75% European ancestry based on ADMIXTURE results | 15,854 | 418,652 |
| XWAS final input file | 15,854 | 418,652 |

⬇

| | Males | Females | Both |
|---|---|---|---|
| MSSNG | 2194 | 520 | 2714 |
| SSC | 1645 | 256 | 1901 |
| SPARK | 1800 | 458 | 2258 |
| HostSeq | 2691 | 3765 | 6456 |
| MGRB | 1220 | 1305 | 2525 |
| Cases | 5639 | 1234 | 6873 |
| Controls | 3911 | 5070 | 8981 |
| Total | 9550 | 6304 | 15854 |

**Figure S1.** Quality Control specifications. The number of samples and variants remaining after each quality control step for autosomes and the X chromosome are shown on the left and right, respectively.

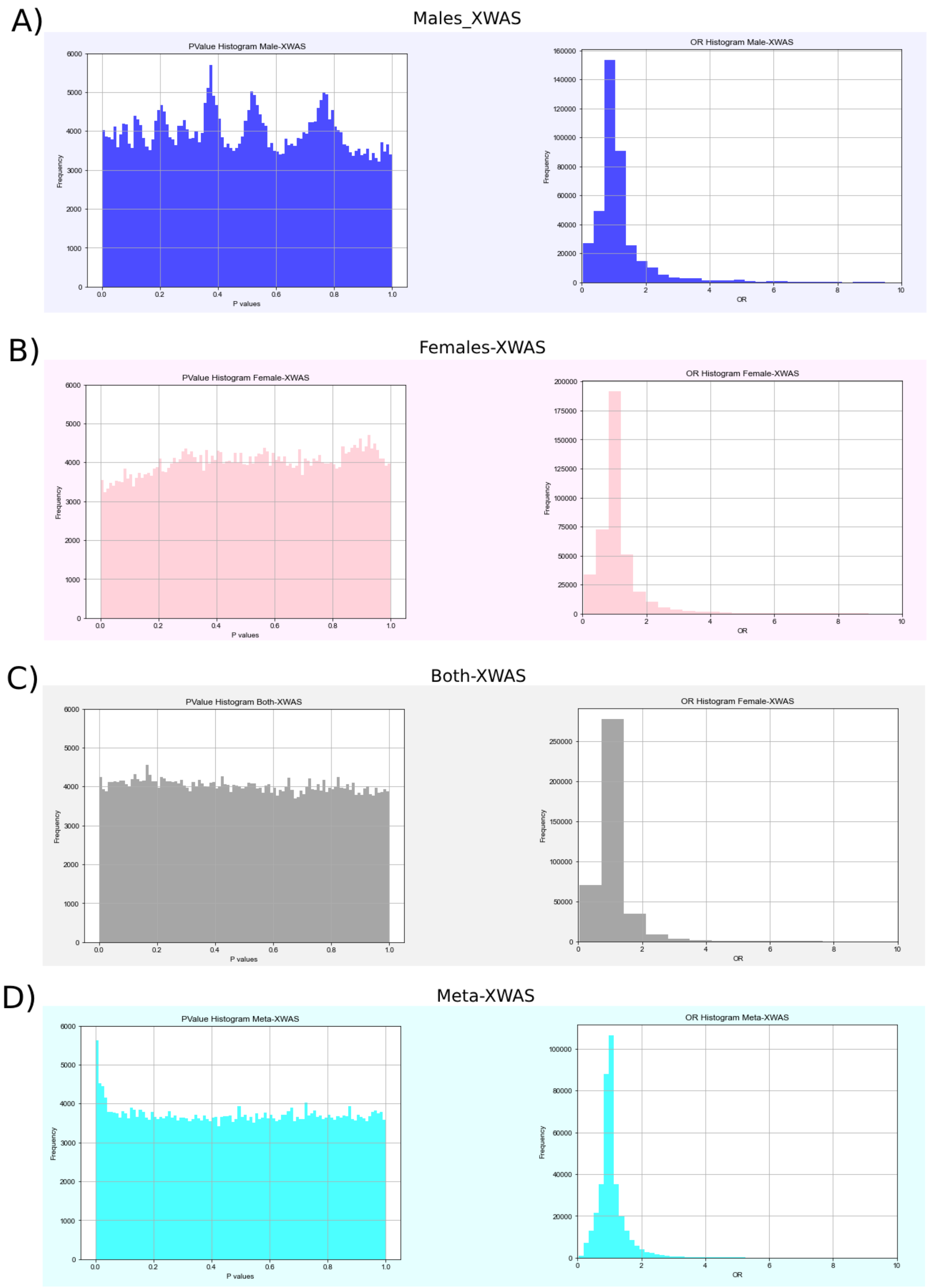


**Figure S2.** P-value and OR Histograms. Each panel shows the p-value histogram (right part) and OR histogram (left part) resultant from our XWAS analysis for (A) Males-XWAS, B)

Females-XWAS, C) Both-XWAS, and D) the Meta-XWAS, a meta-analysis from the sex stratified approaches implemented on GWAMA.

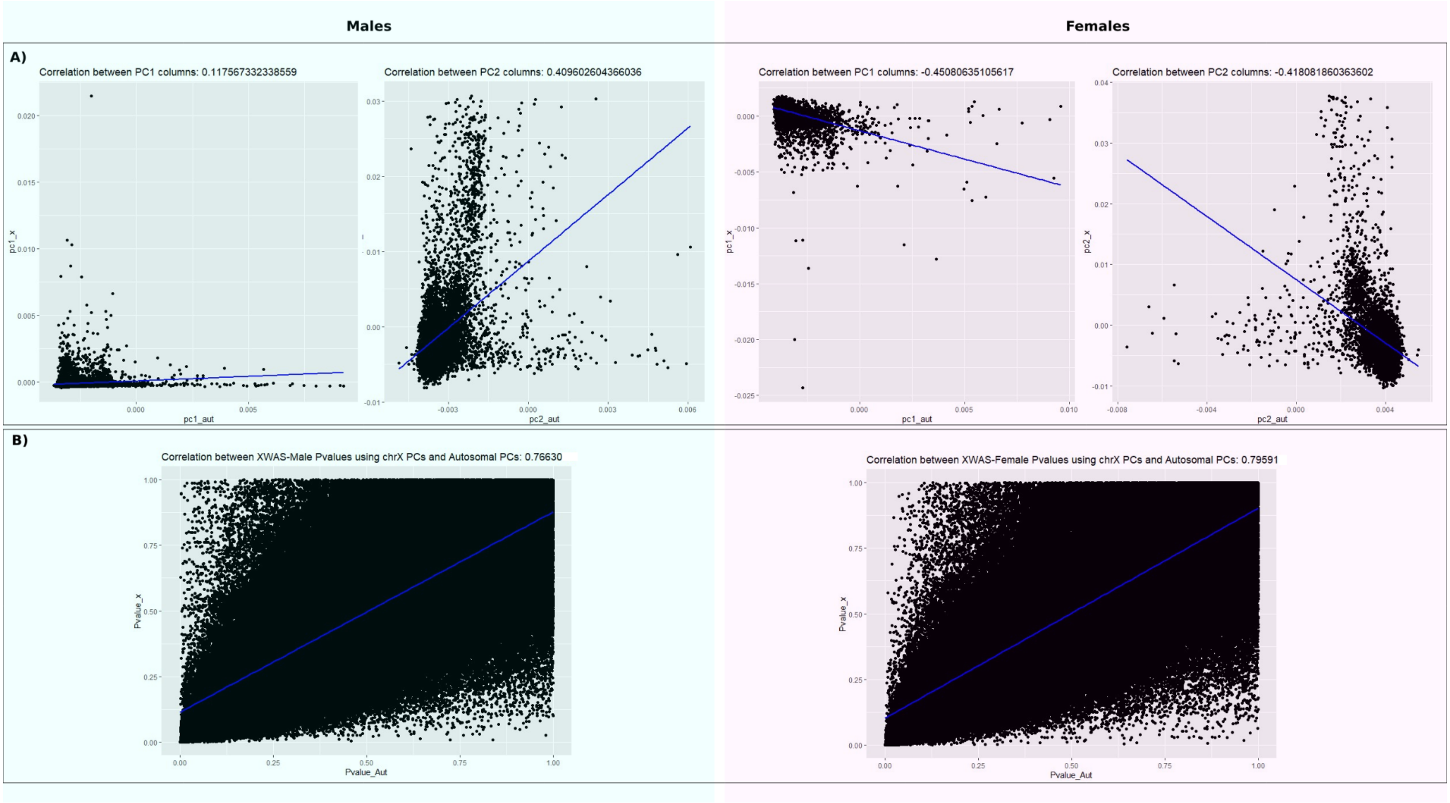


**Figure S3.** Correlation between XWAS results using PCs from Autosomal and X chromosomal data. A) Correlation between PC1 and PC2 from Autosomal chromosomes on the horizontal axis and X chromosome in the vertical axis, in blue from males and in pink from females. B) Correlation between p-value results coming from XWAS using Autosomal data for PCs inference on the horizontal axis and XWAS using X chromosome data for PCs inference on the vertical axis.

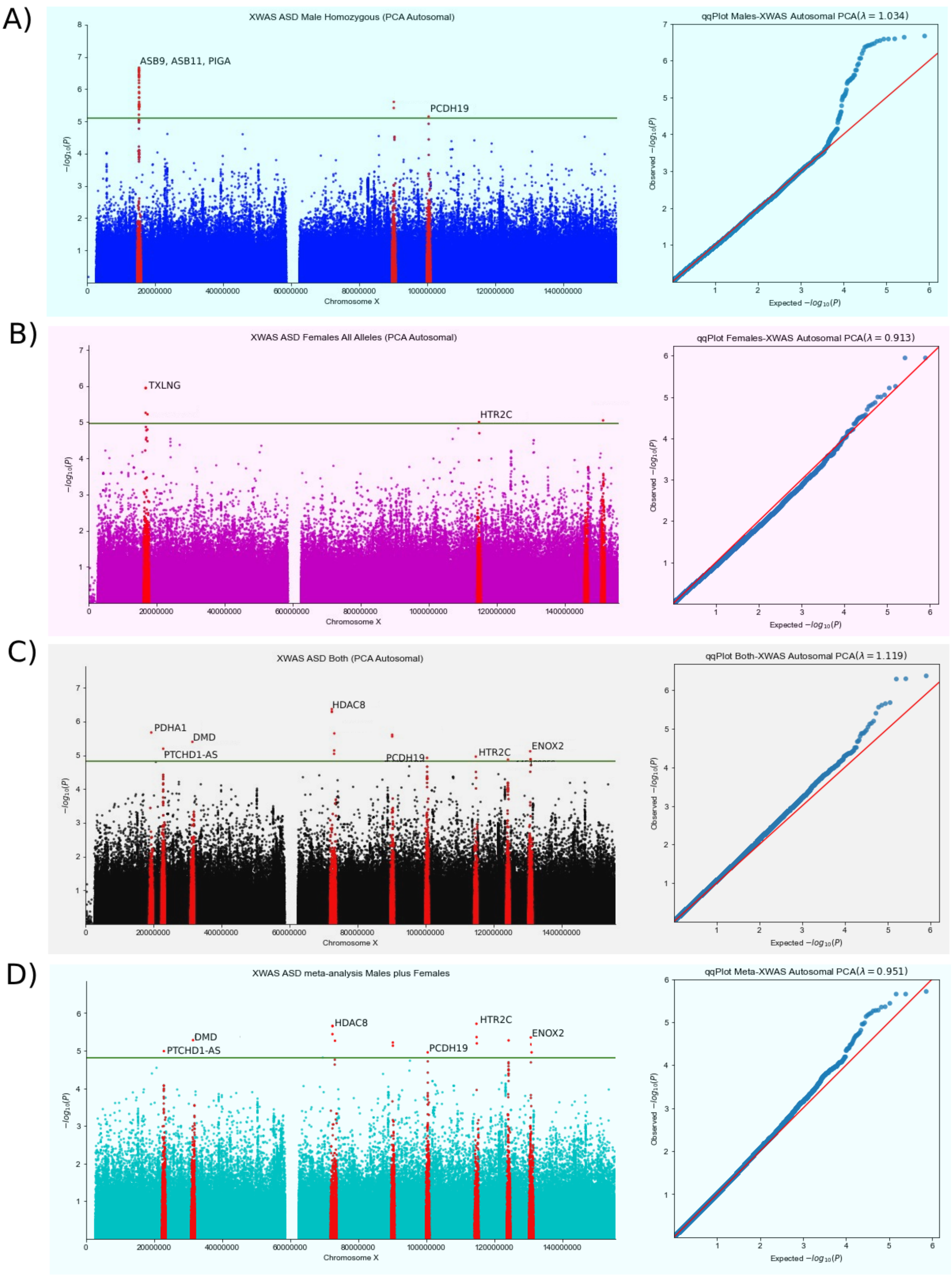


**Figure S4.** Each panel shows the Manhattan (right part) and qqPlot (left part) resultant from our XWAS analysis using the top 10 autosomal Principal Components as covariates (A) Males-

XWAS, B) Females-XWAS, C) Both-XWAS, and D) the Meta-XWAS, a meta-analysis from the sex stratified approaches implemented on GWAMA.
